# Gompertz law based biological age (GOLD BioAge): a simple and practical measurement of biological aging to capture morbidity and mortality risks

**DOI:** 10.1101/2024.11.14.24317305

**Authors:** Meng Hao, Hui Zhang, Jingyi Wu, Xiangnan Li, Yaqi Huang, Meijia Wang, Shuming Wang, Jiaofeng Wang, Jie Chen, Zhi jun Bao, Li Jin, Xiaofeng Wang, Zixin Hu, Shuai Jiang, Yi Li

**Affiliations:** Department of Geriatric Medicine, Huadong Hospital, Shanghai Medical College, Fudan University, Human Phenome Institute, Fudan University, Shanghai, China; State Key Laboratory of Genetic Engineering, Collaborative Innovation Center for Genetics and Development, School of Life Sciences, Fudan University, Shanghai, China; Fudan Zhangjiang Institute, Shanghai, China; School of Global Health, Chinese Center for Tropical Diseases Research, Shanghai Jiao Tong University School of Medicine, Shanghai, 200025, China; Artificial Intelligence Innovation and Incubation Institute, Fudan University, Shanghai, 200438, China; Department of Gerontology, Huadong Hospital, Shanghai Medical College, Fudan University, Shanghai, China; Department of Vascular Surgery, Shanghai Key Laboratory of Vascular Lesion Regulation and Remodeling, Shanghai Pudong Hospital, Fudan University Pudong Medical Center, Shanghai, 201399, China

## Abstract

Biological age reflects actual aging and overall health, but current aging clocks are often complex and difficult to interpret, limiting their clinical application. In this study, we introduced a Gompertz law-based biological age (GOLD BioAge) model that simplified aging assessment. We estimated GOLD BioAge using clinical biomarkers and found significant associations of the difference from chronological age (BioAgeDiff) with risks of morbidity and mortality in NHANES. Moreover, we developed GOLD ProtAge and MetAge using proteomics and metabolomics data, which outperformed the clinical-only model in predicting mortality and chronic disease risks in UK Biobank. Benchmark analysis illustrated that our models exceeded common aging clocks in predicting mortality across diverse age groups in both NHANES and UK Biobank. The results demonstrated that the GOLD BioAge algorithm effectively applied to both clinical and omics data, showing excellent performance in predicting age-related outcomes. Additionally, we created a simplified version called the Light BioAge, which used three biomarkers for aging assessment. The Light model reliably captured mortality risks in three validation cohorts (CHARLS, RuLAS, CLHLS). It significantly predicted the onset of frailty, stratified frail individuals, and collectively identified individuals at high risk of mortality. In summary, the algorithm of GOLD BioAge could provide a valuable framework for aging assessment in public health and clinical practice.

**Highlights:**

1. The algorithm of Gompertz law based biological age (GOLD BioAge) was proposed to construct biological aging clocks with convenient and interpretable calculations, which had better performance in predicting mortality risks.
2. Our approach was applicable to proteomics and metabolomics, yielding ProtAge and MetAge with great clinical prospect to improve accuracy of aging assessment and prevent age-related diseases.
3. The Light BioAge, a simplified version, was developed using age and three biomarkers, and it independently predicted mortality in three cohorts.
4. The Light BioAgeDiff significantly predicted the onset of frailty, stratified frail individuals, and collectively identified individuals at high risk of mortality.

## Introduction

The human aging manifests as progressive physiological changes, physical and cognitive function decline, leading to an increased risk of mortality^1^. There is significant heterogeneity among individuals during aging process^2^, while chronological age may not accurately reflect the actual pace of aging. In addition, aging is the greatest risk factor for most chronic diseases, suggesting that targeting aging itself has the potential to delay multiple aging-associated disease processes^3^. Consequently, aging assessments and treatments have the potential to forecast and prevent functional decline and age-related chronic disease^4^. Some routine clinical biomarkers serve as biomarkers for aging, predicting the risks of functional decline and mortality after adjusting for chronological age^5^. In addition, integrating these biomarkers into composite panels could offer a more comprehensive and powerful assessment of aging compared to single biomarkers alone.

Biological age measures an organism’s biological functioning compared to the expected level for a specific chronological age, reflecting overall health status^6,7^. The Levine’s phenotypic age, which integrated nine biomarkers with chronological age, predicted mortality more accurately than chronological age alone^8^. Building on the concept of phenotypic age, Sheng et al. proposed PCAge to estimate biological age through linear dimensionality reduction; however they could be sensitive to outliers and thresholding effects^9^. Correspondingly, Wei et al presented ENABLAge, integrating machine-learning models with explainable artificial intelligence to ensure high prediction accuracy^10^. In addition to clinical aging clocks, omics-based aging clocks hold significant promises, as they capture more precise dynamic molecular interactions and pathways closely tied to the biological aging process^11^. Specially, the epigenetic biomarkers have been extensively utilized in the DNA methylation aging clocks, such as the Horvath Clock^12^ and GrimAge Clock^13^. Furthermore, multi-tissue aging clocks provide insights into how complex organisms undergo molecular changes with age, offering more detailed information about aging and disease states^11,14,15^. Recently, emerged proteomics and metabolomics data of large cohorts accelerated the development of plasma proteomic and metabolomic aging clocks^16–19^. These proteomics aging clocks showed promising accuracy in predicting mortality and multimorbidity^14,16^. Although these biological aging clocks had excellent performance in forcasting diseases and mortality, the clinical translation remained limited, due to the gap between the scientific research and its application in clinical translational settings^20^.

The complexity of aging clock models, along with issues of interpretability, required features, and generalization capacity, may impede their clinical application. For instance, while DNA methylation clocks are the widely used, biosamples collection and high-throughput sequencing can be both time-consuming and expensive. Therefore, the development of aging clocks should emphasize delivering clinically actionable insights while ensuring affordability, accessibility, and robustness across diverse populations. To tackle these challenges in clinical practice, it is essential to create a computational algorithm that can calculate simplified, robust, and practical biological aging clocks using a small number of effective biomarkers.

The Gompertz law is one of the most widely used mathematical model for describing mortality, and it effectively captures the exponential increase in mortality hazard across adult ages, which strongly fits with empirical mortality data^21^. Also, the model’s simplicity and flexibility allow it to be applied across wide ranges. For example, the Levine’s phenotypic age was proposed based on 10-years mortality risks using the Gompertz model^8^. Also, Kuo at al. proposed proteomic aging clock using proteomics data based on cumulative mortality risks of Gompertz model^19^. Therefore, the Gompertz model provided as a theoretical basis to optimize the phenotypic age for clinical practices.

Here, we developed an algorithm framework for Gompertz law based biological age (GOLD BioAge). The GOLD BioAge constructed aging clocks with a linear combination of chronological age and biomarkers, and linked the its difference from chronological age to morbidity and mortality risks. Then, we applied the GOLD BioAge algorithm on metabolomics and proteomics data in the UKB, to investigate the algorithm validity on omics-based data. Moreover, we compared its prediction performances of mortality with common aging clocks using data from the National Health and Nutrition Examination Survey (NHANES) and UK Biobank (UKB). Finally, we refined and simplified GOLD BioAge as a Light model and validated it across three independent Chinese cohorts: the China Health and Retirement Longitudinal Study (CHARLS), the Chinese Longitudinal Healthy Longevity Survey (CLHLS), and the Rugao Longevity and Ageing Study (RuLAS).

## Results

### Definition and development the GOLD BioAge model

The biological age referred to the age that accurately reflected an individual’s risk of mortality; a higher mortality risk corresponded to an older biological age. Based on the Gompertz law model, we linked chronological age and biomarkers to mortality hazard with the exponential distribution (**Figure 1A**). Consequently, the Gompertz law based biological age (GOLD BioAge) was estimated as the age that aligned with the joint mortality hazard derived from both chronological age and biomarkers. Thus, the Gold Biological Age (GOLD BioAge) was calculated as a linear combination of chronological age and biomarkers.

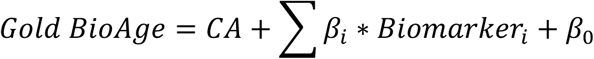

**Figure 1.**
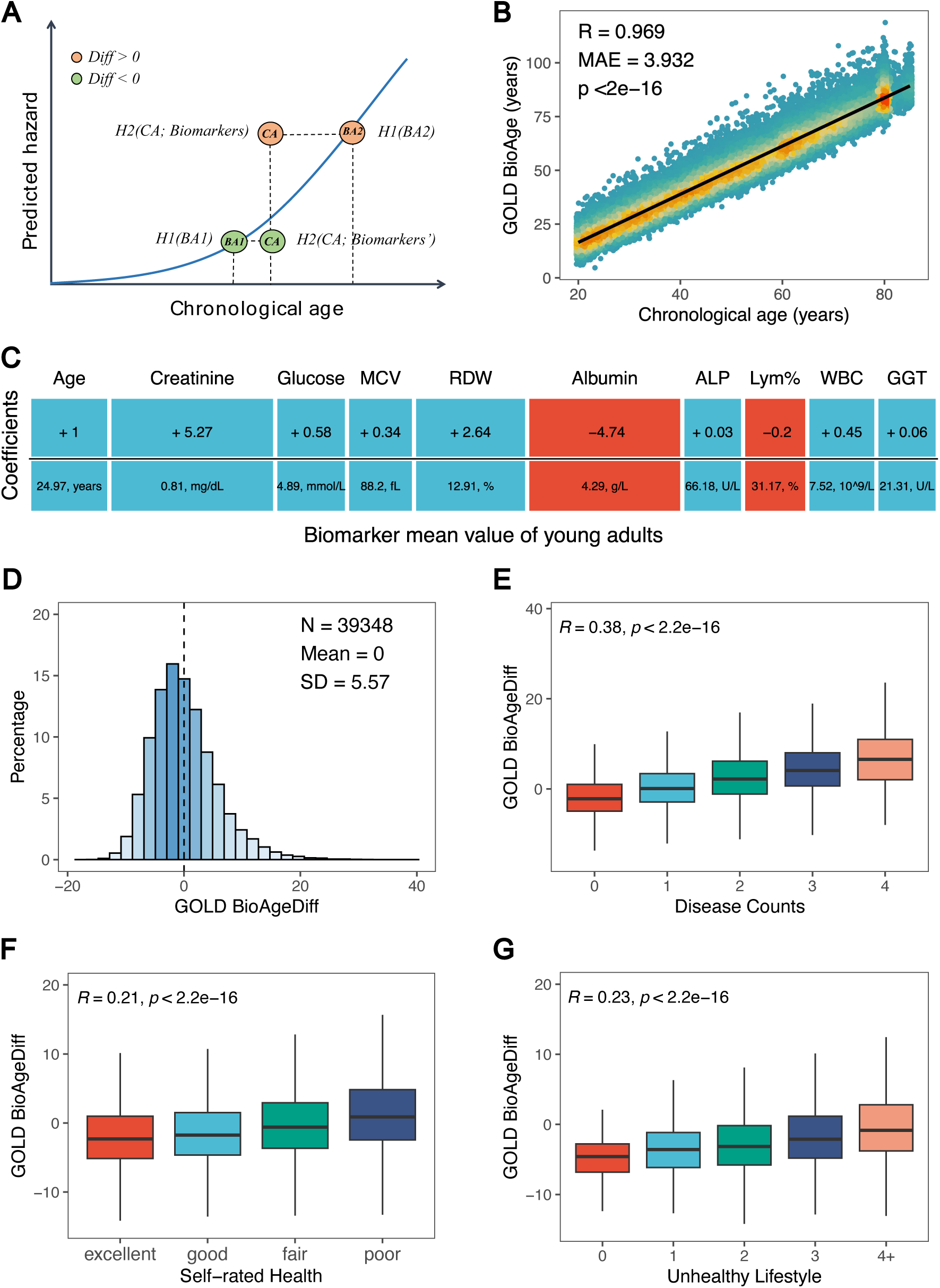
GOLD BioAge and its Association with Health-Related Factors. The diagram (A) illustrated the exponential relationship between mortality hazard and biological age (BA), chronological age (CA). The “Diff” referred to the difference between GOLD BioAge and CA, termed GOLD BioAgeDiff. The scatter plot (B) showed the strong correlation between GOLD BioAge (estimated biological age) and CA. The estimated coefficients for CA and biomarkers (C), used to calculate GOLD BioAge, were displayed, with the mean biomarker values of young adults serving as the reference. The distribution of GOLD BioAgeDiff in NHANES (D). The correlations of GOLD BioAgeDiff with counts of age-related chronic diseases (E), self-rated health (F), and unhealthy lifestyles (G).

In the NHANES, 39,348 samples (49.5 ± 18.0 years old) with CBC and bioassay biomarkers were enrolled in the analysis. After feature selection implemented in LASSO Cox regression (**Figure S1**), we developed a clinical aging clock based on 10 biomarkers, referred as GOLD BioAge, which showed a strong correlation with chronological age (R = 0.969, **Figure 1B**). The GOLD BioAge was the linear combination of chronological age, red blood cell distribution width (RDW), albumin (ALB), creatinine, and etc (**Figure 1C**). This model provided an intuitive interpretation of how biomarker values relate to biological age.

### GOLD BioAgeDiff as a novel aging metric

We then introduced GOLD Biological Age Difference (BioAgeDiff) as the difference between the BioAge and chronological age, to estimate the magnitude of how individuals’ biological age deviated from their chronological age (**Figure 1A, S2**). If the BioAgeDiff was greater/lower than 0, it meant that the person was older/younger than the CA. The BioAgeDiff, as the linear combination of biomarkers, established a clear relationship between changes in biomarkers and shifts in biological age. This calculation of BioAgeDiff made it easy to understand how deviations in biomarkers from reference values affect biological age. For instance, if an individual’s blood glucose level increased by 1 mmol/L, the BioAge would rise by 0.58 years (**Figure 1C**).

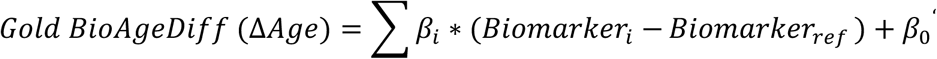

Figure 1D illustrated the distribution of the BioAgeDiff, which was close to the normal distribution (Mean: 0, SD: 5.707). By counting the major chronic diseases, participants with comorbidity had higher BioAgeDiff compared to those without any diseases (Figure 1E). Notably, individuals with four diseases were approximately 5 years older in BioAge. Considering health status, a higher BioAgeDiff was found to be cross-sectionally associated with poorer self-rated health (Figure 1F). Additionally, unhealthy lifestyles, such as smoke and alcohol use, were associated with a higher BioAgeDiff (Figure 1G). These results of the BioAgeDiff were validated in the UKB (**Figure S3**).

The BioAgeDiff was associated with risks of mortality in NHANES and UKB (**Table 1**), with the hazard ratios (HRs) of 1.155 and 1.133, respectively. Survival curve analysis (Figure 2) of 20 years follow-ups revealed that participants in the highest 20% of BioAgeDiff showed a steeper decline in survival probability compared to those in the lowest 20%, especially among middle-aged and older age groups. For instance, individuals aged 65-74 years, about 80% of those in the high-risk group had died after about 16 years, whereas only about 30% of those in the low-risk group had died. The BioAgeDiff could be considered as a measure through linear dimension reduction or projection. Thus, we also compared the performance of BioAgeDiff with those common metrics, including mahalanobis distance statistic^22,23^ (MDS) and principal component analysis^24^ (PCA). In middle-aged (45-64 years) and older age groups (65-85 years), BioAgeDiff outperformed other linear metrics in identifying individuals with high risks of mortality (Figure 2).

**Figure 2.**
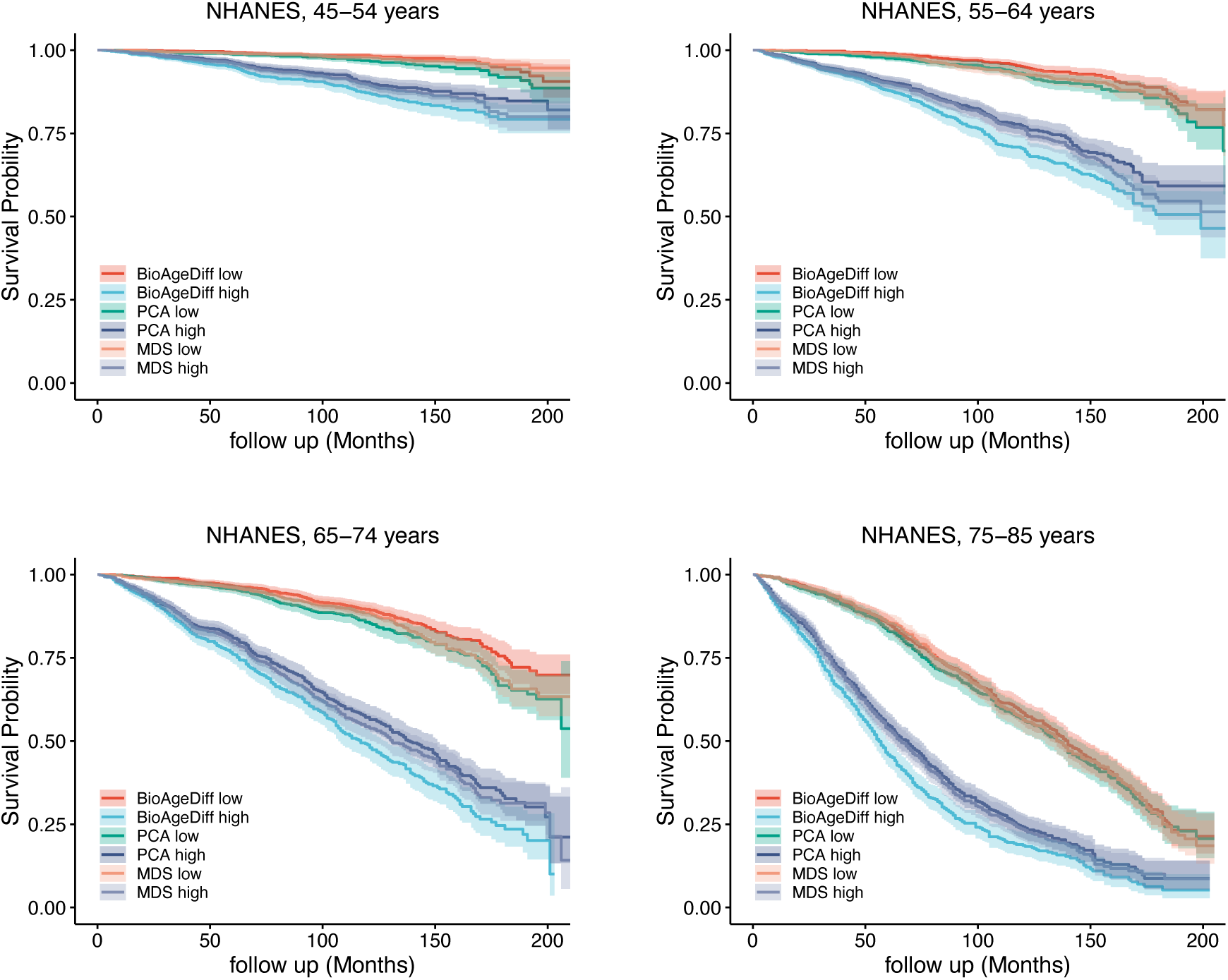
The associations of BioAgeDiff and risks of mortality. The survival plots for individuals categorized by levels of BioAgeDiff, PCA age, and MDS in the NHANES cohort. The high and low groups represent the top and bottom 20% of the age-stratified population (ages 45-54, 55-64, 65-74, and 75-85 years). PCA: Principal Component Analysis; MDS: Mahalanobis Distance Statistics.

**Table 1.** Associations of GOLD BioAgeDiff and its light version with mortality across five cohorts.

| Category/Cohorts | Events/N | Follow-up years | HR (95% CI) | C-index |
| --- | --- | --- | --- | --- |
| **GOLD BioAgeDiff** |  |  |  |  |
| NHANES | 4,716/ 39348 | 8.08 (4.42 - 11.92) | 1.155 (1.150-1.159) | 0.771 |
| UKB | 36,589/ 417,067 | 13.98 (13.21 - 14.72) | 1.133 (1.131-1.135) | 0.677 |
| **Light BioAgeDiff** |  |  |  |  |
| NHANES | 6,373/38,001 | 11.08 (4.33 - 15.33) | 1.197 (1.191-1.204) | 0.697 |
| UKB | 37,619/ 428,266 | 13.98 (13.21 - 14.71) | 1.143 (1.140-1.147) | 0.622 |
| CHARLS | 1,752/17,163 | 9.00 (5.00 - 9.00) | 1.130 (1.118-1.143) | 0.643 |
| RuLAS | 186/ 1,785 | 4.00 (4.00 - 4.00) | 1.093 (1.052-1.136) | 0.585 |
| CLHLS | 813/2,499 | 4.08 (2.54 - 4.08) | 1.060 (1.045-1.075) | 0.575 |

HR: hazard ratio; CI: confidence interval; All cox proportional hazard regression models were adjusted for age and sex. The C-index was calculated in the crude model. Follow-up years were shown with the median (interquartile range).

### Application of GOLD BioAge on metabolomics and proteomics

To further investigate the utility of GOLD BioAge with multi-omics biomarkers, we applied our algorithm to create the MetAge and ProtAge models based on blood NMR metabolomics and proteomics data in the UKB, respectively. Like the clinical-based BioAge, the omics-based aging clocks showed strong correlations with chronological age and age-related factors (Figure 3A**, S4**). The ProtAge exhibited significant abilities to capture mortality risks, surpassing MetAge, clinical BioAge and chronological age (Figure 3B). For all-cause mortality, ProtAge achieved a C-index of 0.790, while MetAge and BioAge reached 0.747 and 0.738, respectively. Additionally, these results were consistent across different age groups and causes-specific mortality (**Table S11**). Notably, among young adults (<45 years), ProtAge demonstrated a C-index of 0.793 in survival analysis, highlighting its effectiveness in predicting premature mortality risk (Figure 3C). For specific mortality, ProtAge recorded a C-index of 0.754 for cancer mortality and 0.850 for heart disease mortality, the highest among the three aging clocks. Individuals in the top 20% of ProtAgeDiff exhibited the highest cumulative mortality incidence rates throughout the follow-up, compared to those with MetAgeDiff and BioAgeDiff (**Figure S5**). These findings further emphasized the superiority of proteomic biomarkers over metabolomics and clinical biomarkers in predicting mortality.

**Figure 3.**
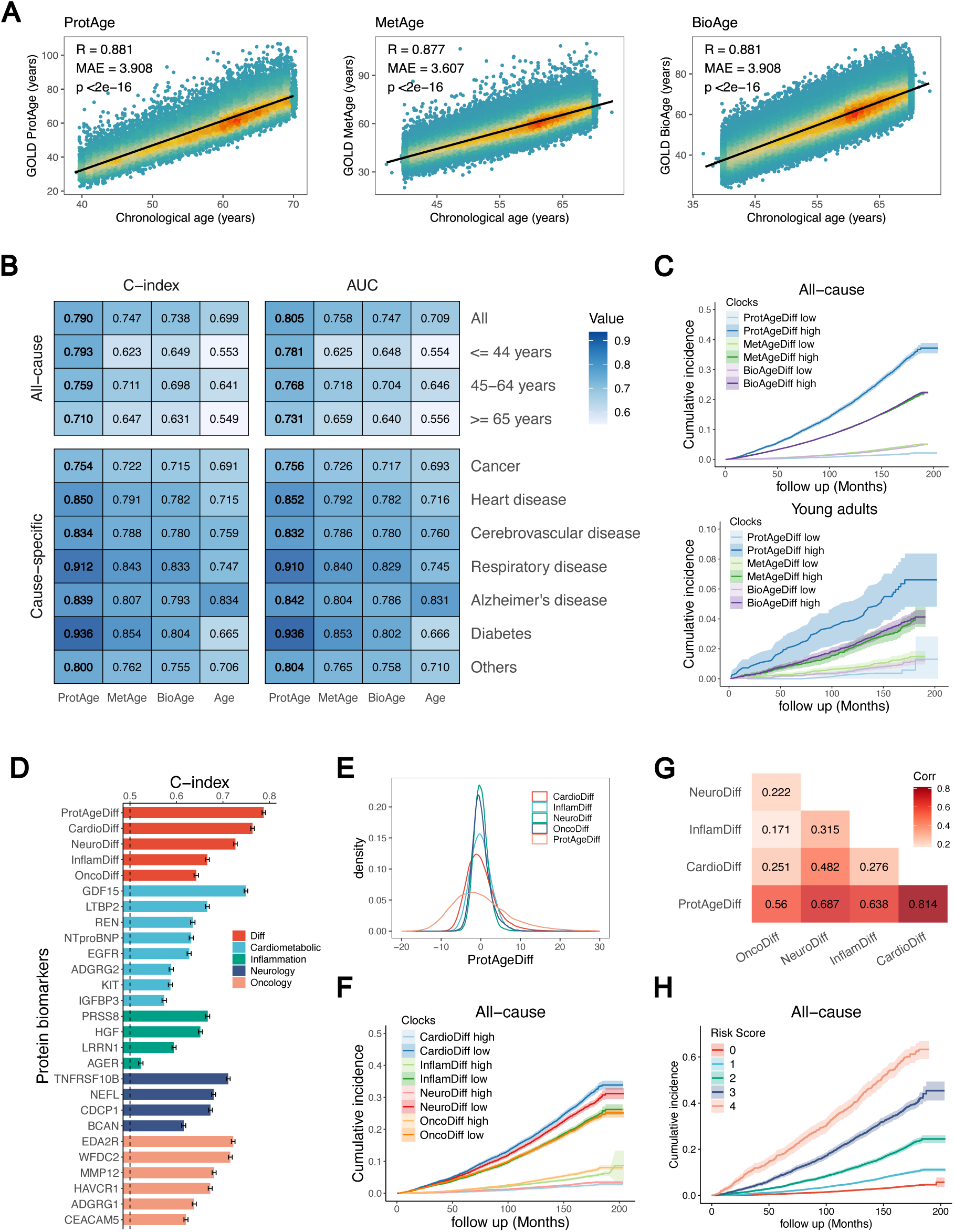
The associations of GOLD ProtAge, MetAge, and BioAge with mortality in UK Biobank. (A) Correlations between the three aging clocks and chronological age. (B) C-index values from survival analysis and the AUC for 10-year mortality prediction, comparing the three aging clocks and chronological age, with results for all-cause (age-stratified) and cause-specific mortality. (C) Survival curves for individuals classified by ProtAgeDiff, MetAgeDiff, and BioAgeDiff in the general population (top panel) and young adults (bottom panel, <45 years old). High and low risk groups were defined as the top and bottom 20% of the population. ProtAgeDiff, MetAgeDiff, and BioAgeDiff represented the differences between ProtAge, MetAge, and BioAge and chronological age, respectively. The C-index for ProtAgeDiff and its subpanels and proteins were shown. ProtAgeDiff consisted of CardioDiff, InfamDiff, NeuroDiff, and OncoDiff, which were linear combinations of cardiometabolic, inflammatory, neurological, and oncological proteins. (E) Density plots and (G) a correlation heatmap (filled with Pearson correlation coefficients) of these subpanels were presented. (F) Survival plots based on ProtAgeDiff subpanels and (H) the risk score, which was the count of high-risk factors derived from ProtAgeDiff subpanels.

Then, we decomposed the ProtAgeDiff into contributions from cardiometabolic (CardioDiff), inflammatory (InflamDiff), neurological (NeuroDiff), and oncological (OncoDiff) proteins (Figure 3D-E), which may reveal various aspects of aging mechanisms. CardioDiff and NeuroDiff emerged as the top two contributors to ProtAgeDiff, demonstrating the highest C-index in survival analysis (Figure 3F**, S6**). Within these proteins (**Table 2**), GDF15, NTproBNP, and EGFR have been identified as aging biomarkers, while NEFL is frequently highlighted among neurological proteins. Given the relative independence of the four ProtAgeDiff categories (Figure 3G), we took the counts within the high-risk group (top 20% of Cardio/Neuro/Inflamm/Onco Diff) into a risk score ranging from 0 to 4. This risk score effectively identified individuals at high risk of mortality (Figure 3H); for example, over 60% of those scoring 4 had died within approximately 16 years due to all-cause mortality. In summary, ProtAge and its ProtAgeDiff serve as exceptional aging clocks for predicting mortality risk, and the ProtAgeDiff calculation allows us to analyze the aging process across four distinct biological categories.

**Table 2.**
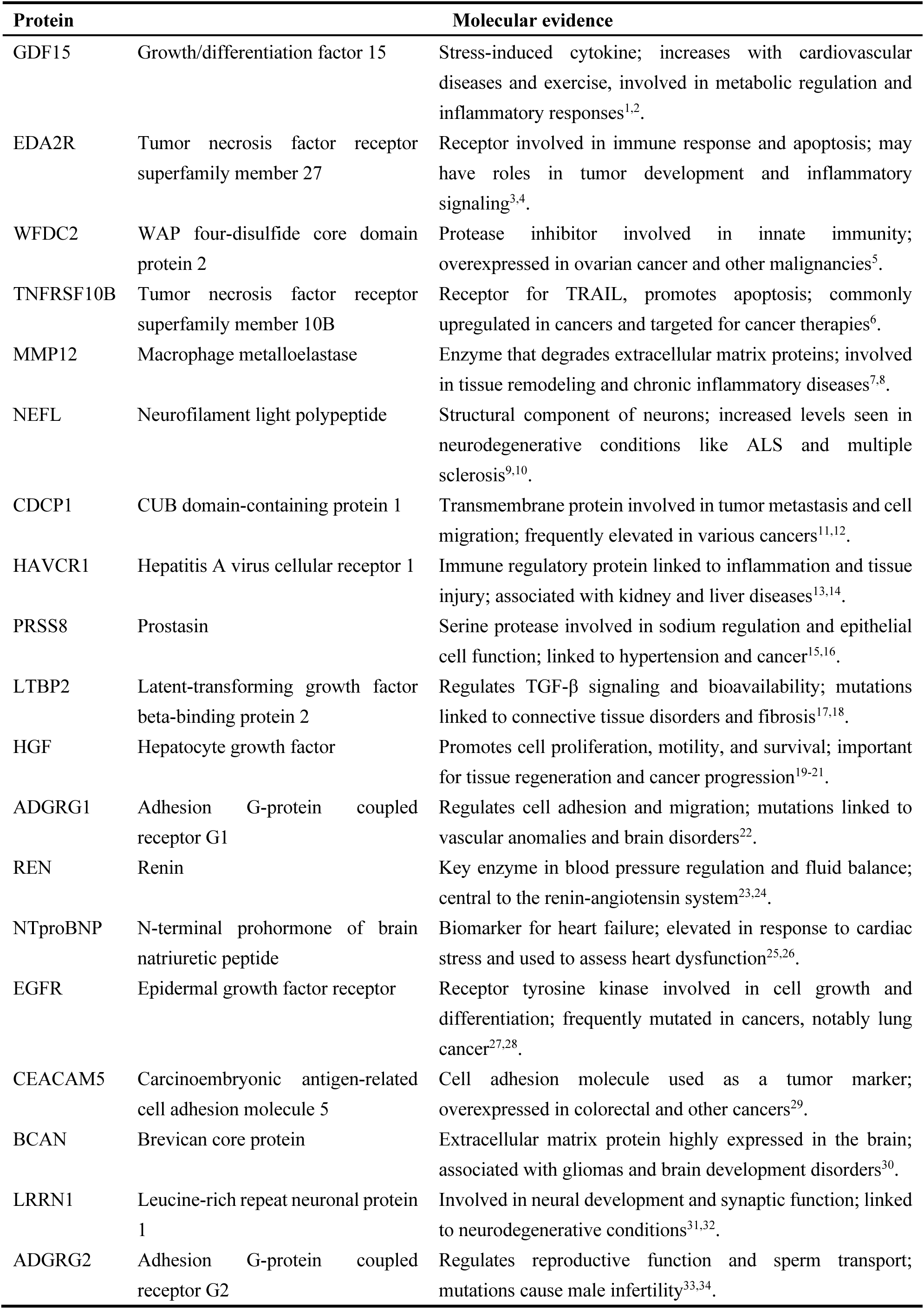
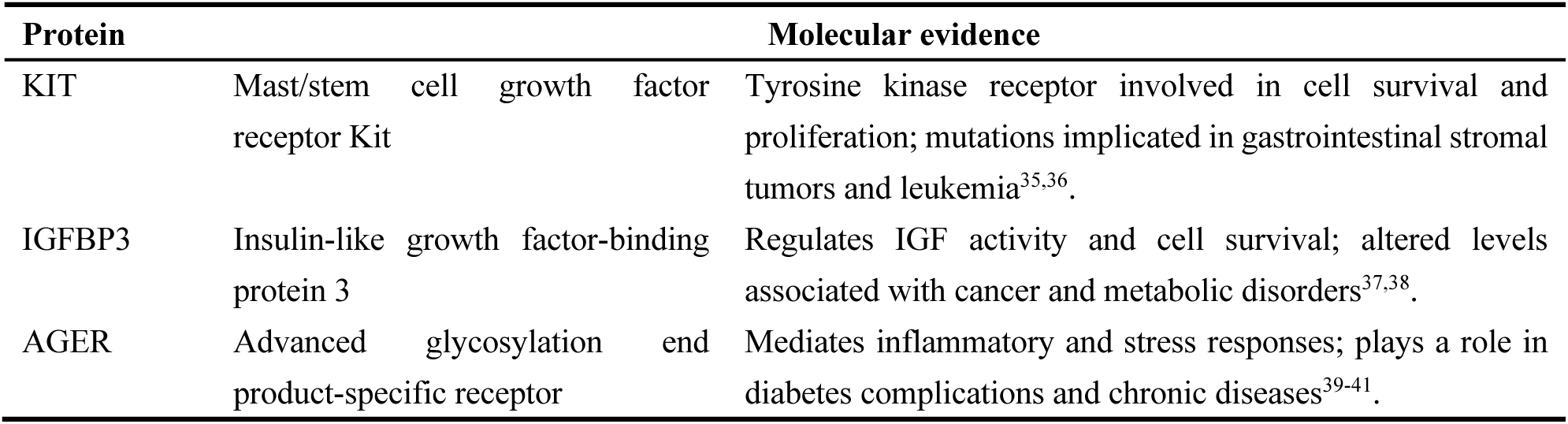
Biological curation of ProtAge-related proteins.

### GOLD BioAge and incident chronic diseases

To investigate the potential of GOLD BioAge in predicting the incidence of common chronic diseases, we included cancer, myocardial infarction, heart failure, stroke, chronic obstructive pulmonary disease (COPD), and dementia in the association analysis. The cox proportional hazards model provided distinct insights into disease risk, showing that one 1-year increase in the biological age was associated with elevated disease risks (Figure 4A). For instance, in the case of cancer, a 1-year increment in ProtAge, MetAge and BioAge was associated with a 2.7%, 1.8%, and 1.6% and increase of hazard ratios (HRs), respectively. This trend was consistent across other diseases, such as myocardial infraction and stroke. Moreover, the ProtAge model demonstrated slightly higher HRs and C-index values for most specific diseases compared to the BioAge model. In dementia, for example, the HR of ProtAge reached 1.078, while BioAge had an HR of 1.051. Similarly, the MetAge model exhibited robust performance across diseases like myocardial infraction and stroke, with HRs of 1.081 and 1.066, respectively. These results highlighted the value of ProtAge and MetAge in predicting incident chronic diseases in large cohorts.

**Figure 4.**
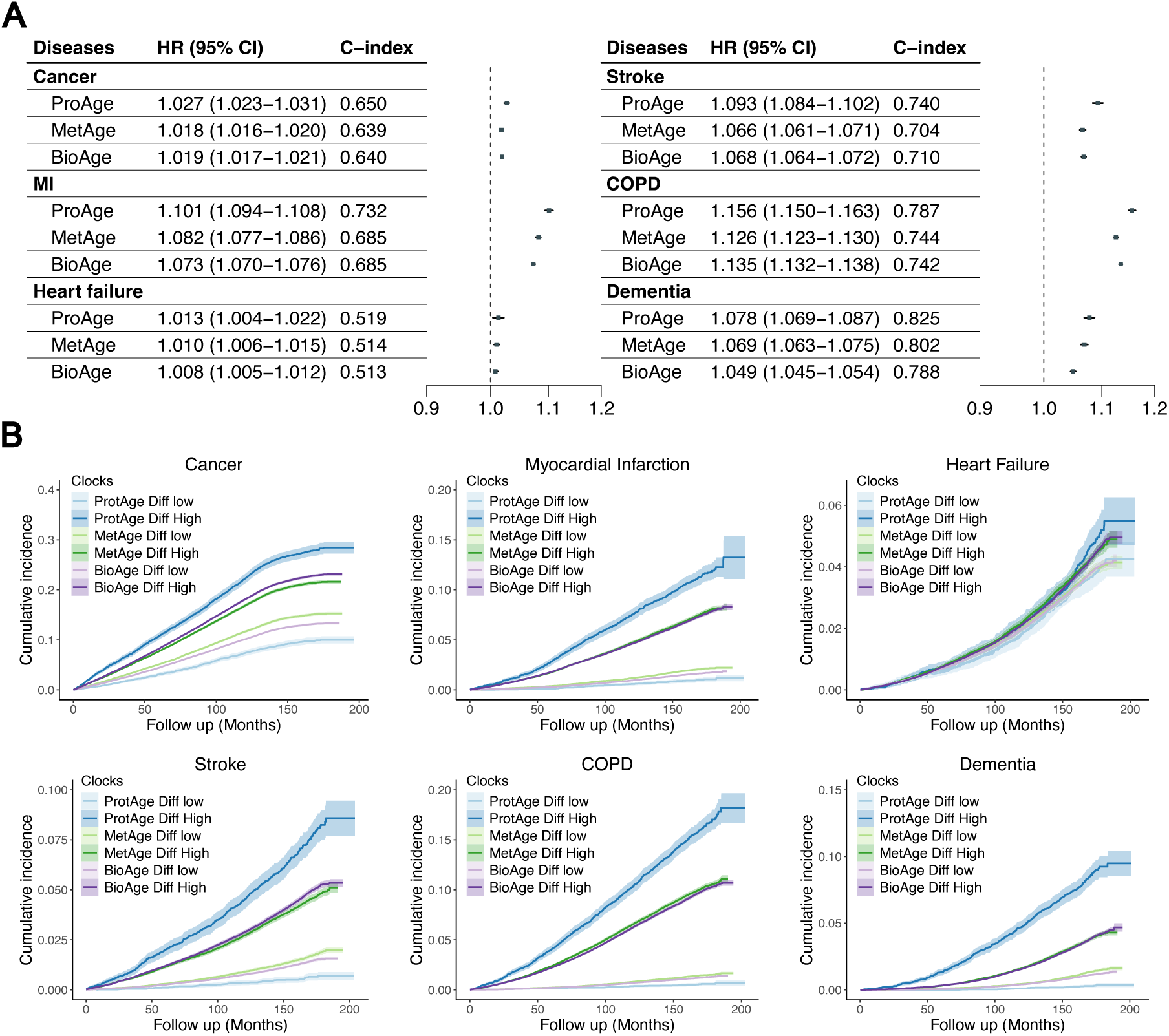
Associations between GOLD ProtAge, MetAge, and BioAge and the incidence of age-related chronic diseases. The forest plots (A) illustrated the hazard ratios and C-index for ProtAge, MetAge, and BioAge across different diseases in the UKB. These associations were adjusted for age and sex. MI: myocardial ischemia; COPD: chronic obstructive pulmonary disease; CI: confidence interval. Survival plots were displayed based on the differences between ProtAge, MetAge, and BioAge relative to chronological age, referred to as ProtAgeDiff, MetAgeDiff, and BioAgeDiff. The high and low groups corresponded to the top and bottom 20% of the population, respectively.

Cumulative disease incidence trajectories were presented based on aging pace, measured by ProtAgeDiff, MetAgeDiff, and BioAgeDiff (Figure 4B). The differences between the highest and lowest ProtAgeDiff groups were most pronounced among the three metrics, indicating that ProtAge was particularly effective in predicting the onset of chronic diseases. Over a follow-up period of 16 years, cumulative mortality rates for cancer, myocardial infarction, heart failure, stroke, chronic obstructive pulmonary disease (COPD), and dementia in the high ProtAgeDiff group were 28.46%, 13.24%, 5.49%, 8.58%, 18.20%, and 9.49%. Overall, these findings underscore the significance of ProtAge and MetAge in forecasting age-relate chronic diseases.

### Comparison with other aging clocks

To investigate the validity of our models, we compared the mortality prediction performance of the GOLD BioAge model with Levine phenotypic age, KDM biological age (KDM-BA), and chronological age in the NHANES (8,106 participants, aged 47.0 ± 16.3 years) and UKB (265,541 participants, aged 56.5 ± 8.0 years). These aging clock models were constructed using clinical biomarkers, with chronological age included as a reference.

Figure 5 showed the C-index of survival analysis and AUC values for 10-year mortality prediction of these aging clocks. The BioAge model significantly showed better overall performance than any other biological and chronological age across the NHANES and UKB dataset, both in the overall sample and within specific age groups. For example, the BioAge model achieved a C-index of 0.847 in the entire cohort, outperforming the Levine’s phenotypic age (0.845), KDM (0.827), and chronological age (0.822). As for cause-specific mortality, the GOLD BioAge model showed the highest the value of C-index and AUC among these aging clocks. For example, for mortality concerning respiratory disease, C-index of the BioAge were 0.885 in NHANES and 0.828 in UKB. Taking the NHANES III as the validation dataset (**Figure S7**), the GOLD BioAge also showed competitive performance, compared with these common aging clocks. These results shown the validity of our biological age algorithm and its efficiency for capture mortality risks.

**Figure 5.**
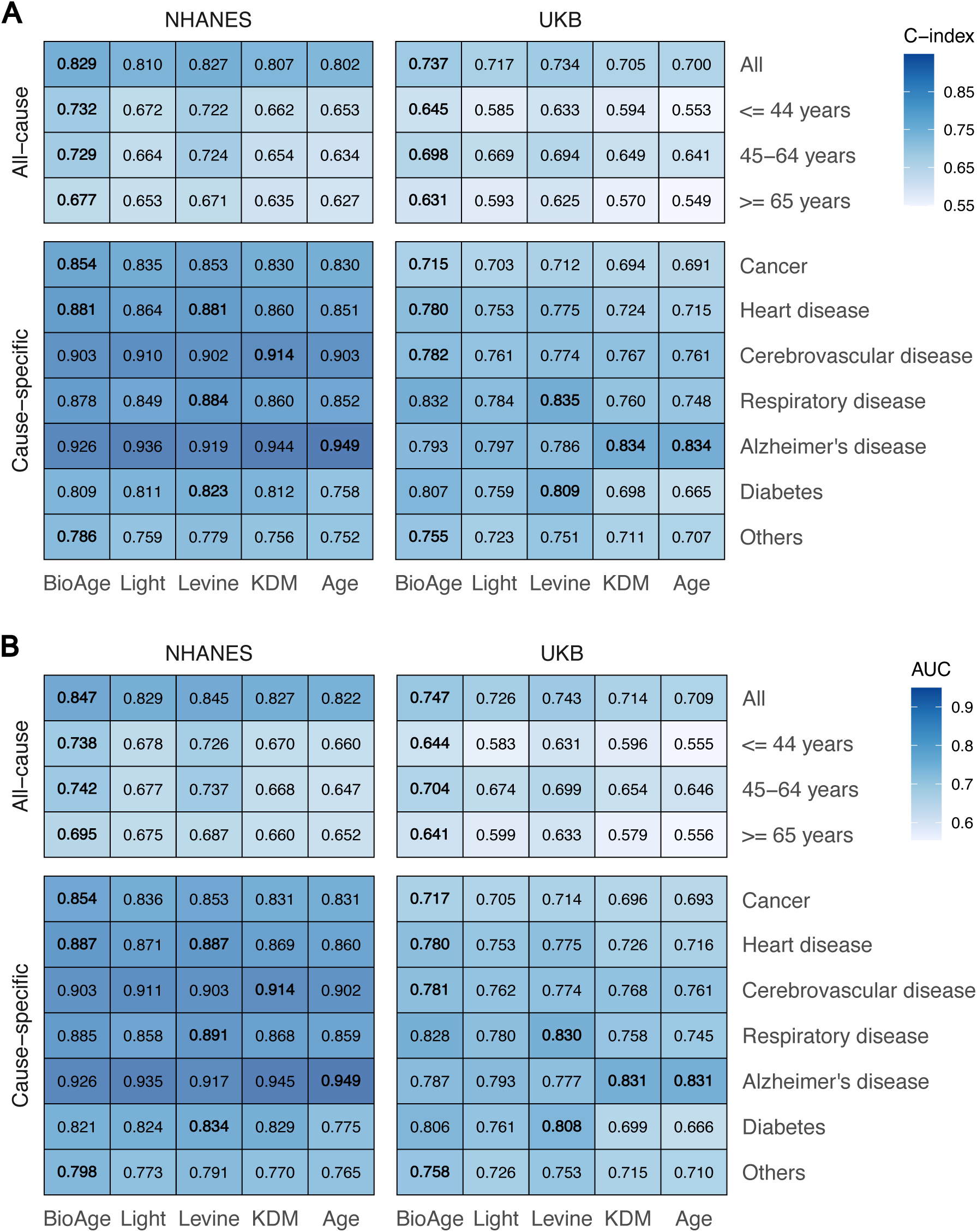
Comparison of GOLD BioAge and other common aging clocks in predicting mortality in NHANES and UKB. The C-index in survival analysis (A) and AUC value of 10-year mortality prediction (B) of these aging clocks were shown. Both all-cause (age-stratified) and cause-specific mortality are considered. The highest value was marked with bold. The BioAge, Light, Levine and KDM referred to the GOLD BioAge, its light version, Levine’s phenotypic age, KDM algorithm derived age, respectively.

### Light BioAge for practice simplicity

For clinical practice simplicity, we refined and simplified GOLD BioAge as a light version called the Light BioAge (**Figure S8**). The Light BioAge model included age, serum creatinine, glucose, and CRP (log-transformed). The calculating formula is as followed:

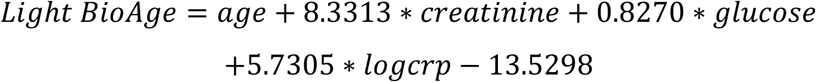

In NHANES, 38,001 samples (49.6 ± 18.3 years old) with these three biomarkers were enrolled. And the Light BioAge was strongly correlated with chronological age (R = 0.987), which accounted for 93.73% of the variance in the GOLD BioAge. Its difference from chronological age (Light BioAgeDiff) was positively correlated with age (Figure 6B), which followed a nearly normal distribution (Figure 6C). It also showed significant associations with comorbility, self-rated health, unhealthy lifestyles, and risks of mortality (Figure 6D-G, **Table 1**).

**Figure 6.**
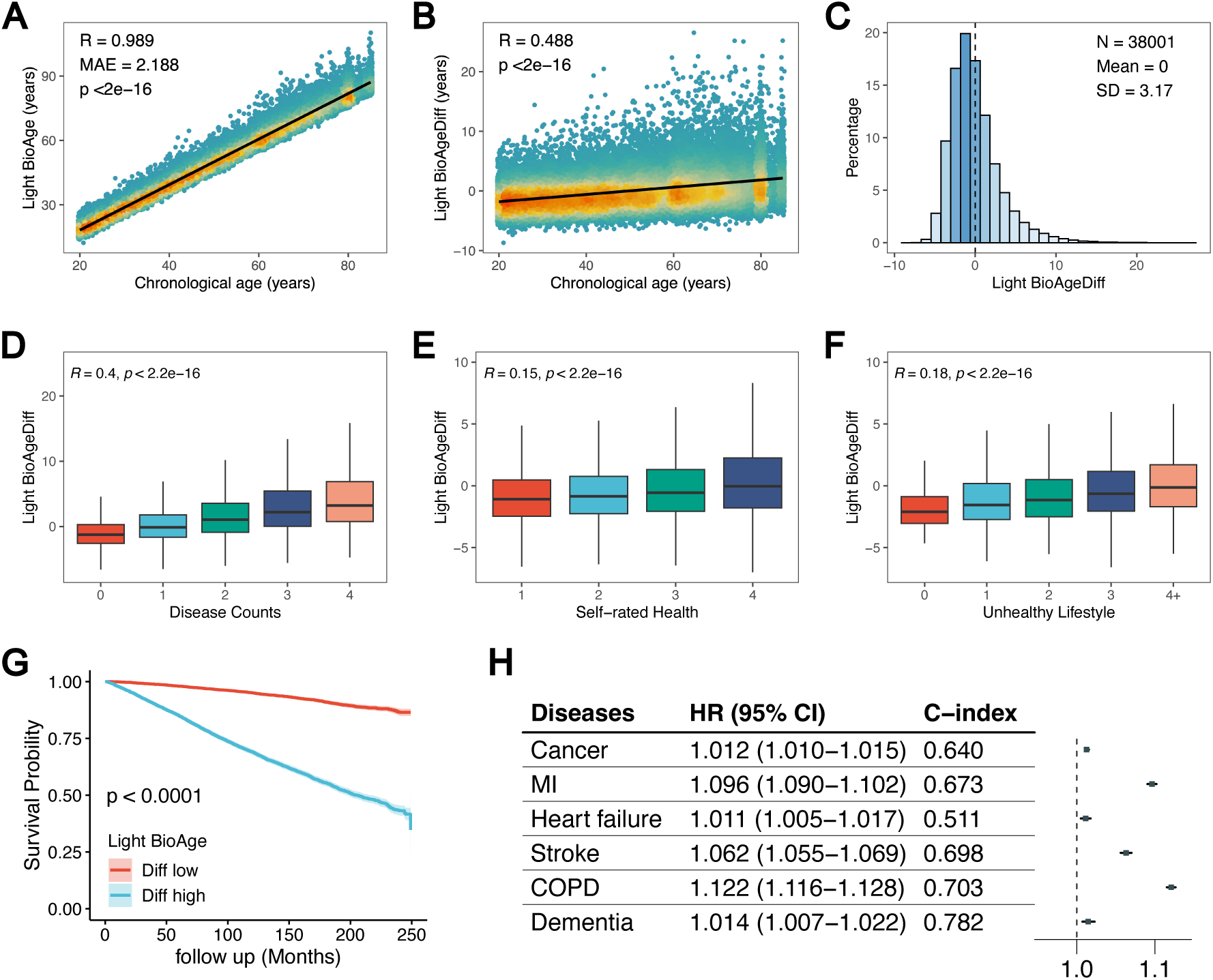
The Light BioAge and its associations with age-related factors and outcomes. The correlation of Light BioAge with age (A). And the difference of Light BioAge with age also correlated with age (B) and its distribution (C). The correlations of Light BioAgeDiff with counts of age-related chronic diseases (D), self-rated health (E), and unhealthy lifestyles (F). The survival plot (G) based on Light BioAgeDiff levels, with the top and bottom 20% of the population representing the high and low groups. The forest plots (H) showed the hazard ratios and C-index of Light BioAge in relation to chronic diseases, adjusted for age and sex. MI: myocardial ischemia; COPD: chronic obstructive pulmonary disease; HR: hazard ratio; CI: confidence interval.

Compared to GOLD BioAge model, the Light BioAge model, utilizing the fewest indicators, demonstrated competitive predictive accuracy (Figure 5). In the NHANES dataset, while the full BioAge model achieved a higher C-index of 0.832 for all-cause mortality, the Light model demonstrated competitive performance with a C-index of 0.811. Further, we found that the C-index of the Light BioAge was very close to previous prominent measures, such as the Levine’s phenotypic age and KDM. For example, for mortality of cerebrovascular disease, the C-index of Light BioAge reached 0.910, comparable to phenotypic age (0.902) and KDM (0.914) in NHANES. Notably, to enhance clinical applicability, we identified its performance in predicting incident chronic disease. For instance, the Light BioAge model demonstrated HR of 1.116, 1.099, and 1.077 for COPD, myocardial infarction, and stroke, respectively (Figure 6H). These results highlighted that the Light BioAge provided a robust and practical alternative while remaining competitive with other aging metrics.

### Light BioAge predicted mortality in validation cohorts

We further validated the Light BioAge in three independent datasets, including the CHARLS (17,163 participants, aged 58.4 ± 10.05 years), RuLAS (1,785 participants, aged 77.0 ± 4.2 years), and CLHLS (2,499 participants, aged 85.5 ± 12.0 years). In the three cohorts (**Table 1**), it documented 1752, 186, 813 deaths during the median follow-up period of 9.0, 4.0, 4.1 years, respectively.

The Light BioAge was strongly correlated with chronological age across the three cohorts (Figure 7A). In the full samples, the Light BioAge achieved AUC values of 0.792 in CHARLS, 0.809 in CLHLS, and 0.746 in RuLAS (Figure 7B). These values were higher than those for chronological age, which were 0.774, 0.790, and 0.646, respectively. Notably, the Light BioAge outperformed chronological age in individuals aged between 60-79 years with AUC exceeding 0.790 in both CLHLS and RuLAS; it also maintained a robust AUC near 0.8 for those aged 75 and older, significantly outperforming chronological age. Participants with high BioAgeDiff (top 20%) experienced a more pronounced decline in survival probability compared to those with low BioAgeDiff (botteom 20%) across CHARLS, RLAS, and CLHLS (Figure 7C). By the end of follow-up periods in each cohort, the survival probabilities of individuals in the high-risk groups were about 75%, 85% and 55%, respectively.

**Figure 7.**
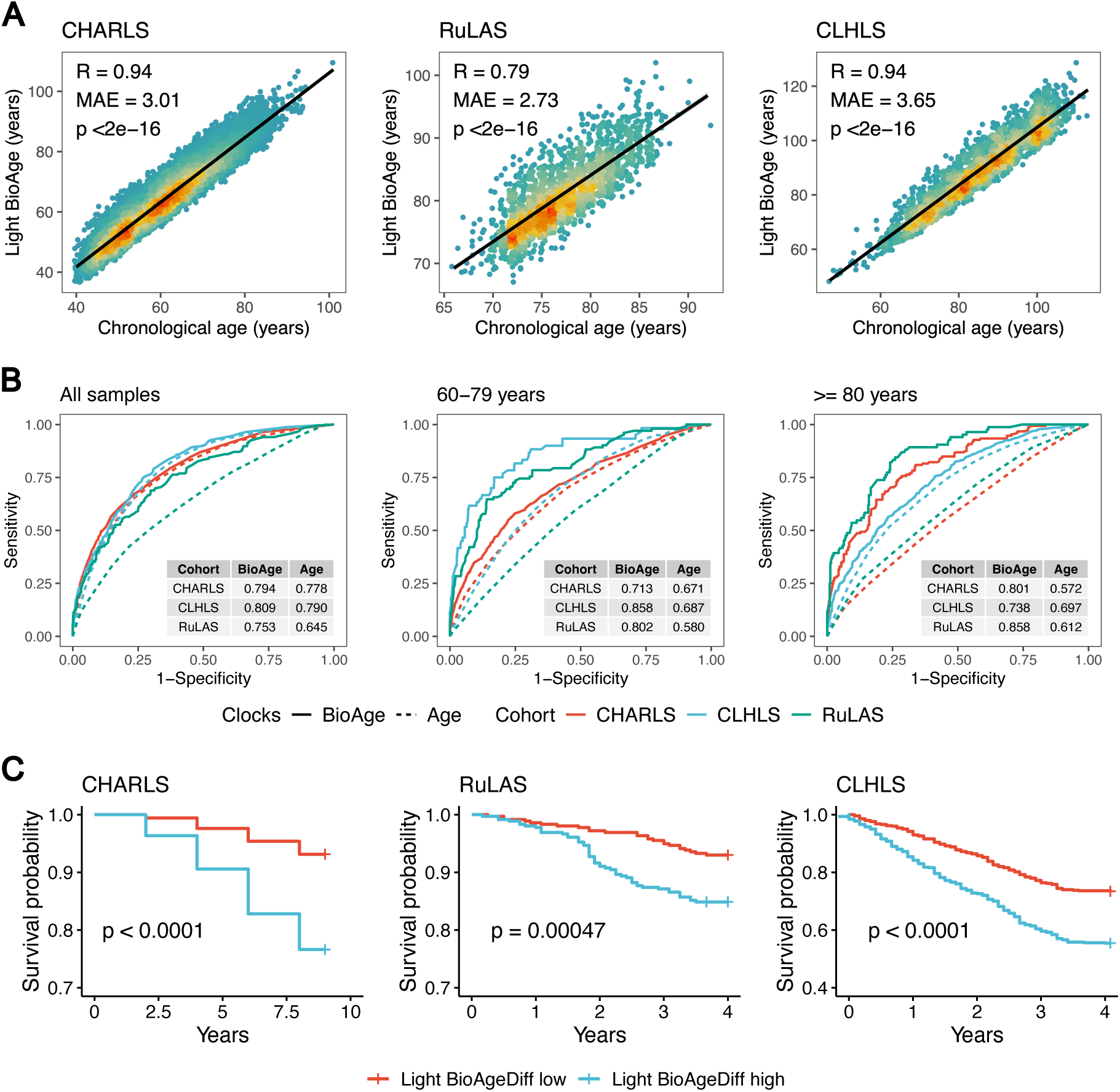
Validations of the Light BioAge in three independent cohorts. The correlations (A) of Light BioAge with age in CHALS, RuLAS and CLHLS. The ROC curves (B) of Light BioAge (solid lines) and age (dotted lines) for predicting mortality across all samples, and within age-stratified groups (<80, >=80 years old). Survival plots (C) depicted mortality trajectories of individuals categorized based on Light BioAgeDiff levels, with the top and bottom 20% represented as high and low groups in CHARLS, RuLAS, and CLHLS.

With human aging as a longitudinal process, we examined the dynamic changes of Light BioAgeDiff between wave 1 and wave 3 of CHARLS (Figure 8A). The Light BioAge in the two waves were strongly correlated (R=0.915, Figure 8B), while the Light BioAgeDiff showed a moderate correlation (R=0.475). According to Light BioAgeDiff, paticipants were classified into slow (Diff<0), normal (0<=Diff<5) and fast (Diff>5) aging groups, subsequently classifying them into seven categories based on their aging status across both waves (Figure 8C). The stable slow-aging groups across the two waves were taken as the reference. Compared with the reference, the stable fast-aging groups and accelerated aging groups (slow/normal to fast) exhibited the highest mortality risks (Figure 8D-E). In addition, the decelerated aged (fast to slow/normal) had reduced mortality risks.

**Figure 8.**
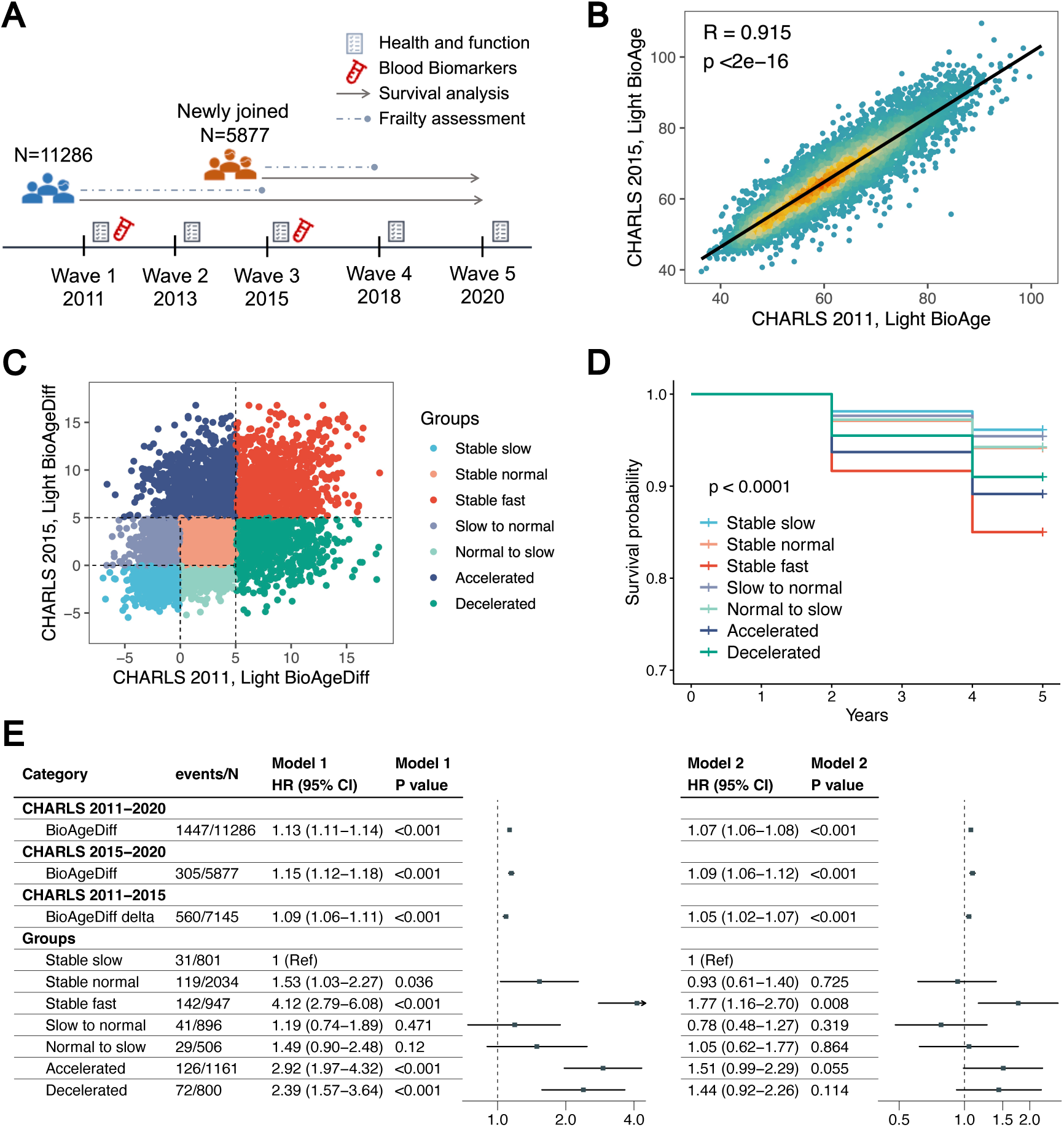
The Light BioAge, its dynamics and mortality in CHARLS. Illustration (A) detailing the study designs across five waves in CHARLS. Correlation (B) between Light BioAge values in wave 1 (2011) and wave 3 (2015). Scatter plot (C) displayed Light BioAgeDiff in wave 1 (2011) and wave 3 (2015), with dotted lines indicating Light BioAgeDiff values of 0 and 5. Individuals were divided into 7 groups based on the changes in Light BioAgeDiff, with survival plots (D) and forest plots (E) provided. Model 1 represented the crude model, while Model 2 adjusted for age and sex.

### Light BioAgeDiff, frailty and mortality risks

Next, we explored the associations of Light BioAgeDiff with frailty as assessed by the frailty index that included age-related chronic diseases, self rated health, basic and instrucmental activities of daily living and mobility capacity. In CHARLS 2011 and 2015, the frailty status were associated with BioAgeDiff, in which the frail individuals were 1.14 and 1.20 years old than the robust counterparts (Figure 9A). During longtidinal follow-ups (2011-2015, 2015-2018), the baseline BioAgeDiff was associated with incident frailty (odds ratio [95% CI]: 1.02 [1.01-1.04]; 1.04 [1.01-1.07], Figure 9B). The paticipants categoried within the fourth quantile of BioAgeDiff had the highest risks. Using the BioAgeDiff as a measure of biologica aging, we examined the mediation role of functional decline, measured by frailty index, in the associations of BioAgeDiff with mortality risks (Figure 9C). The mediation proportion of frailty index was about 26.4% (p<0.001) while its increase accounted for 6.48%.

**Figure 9.**
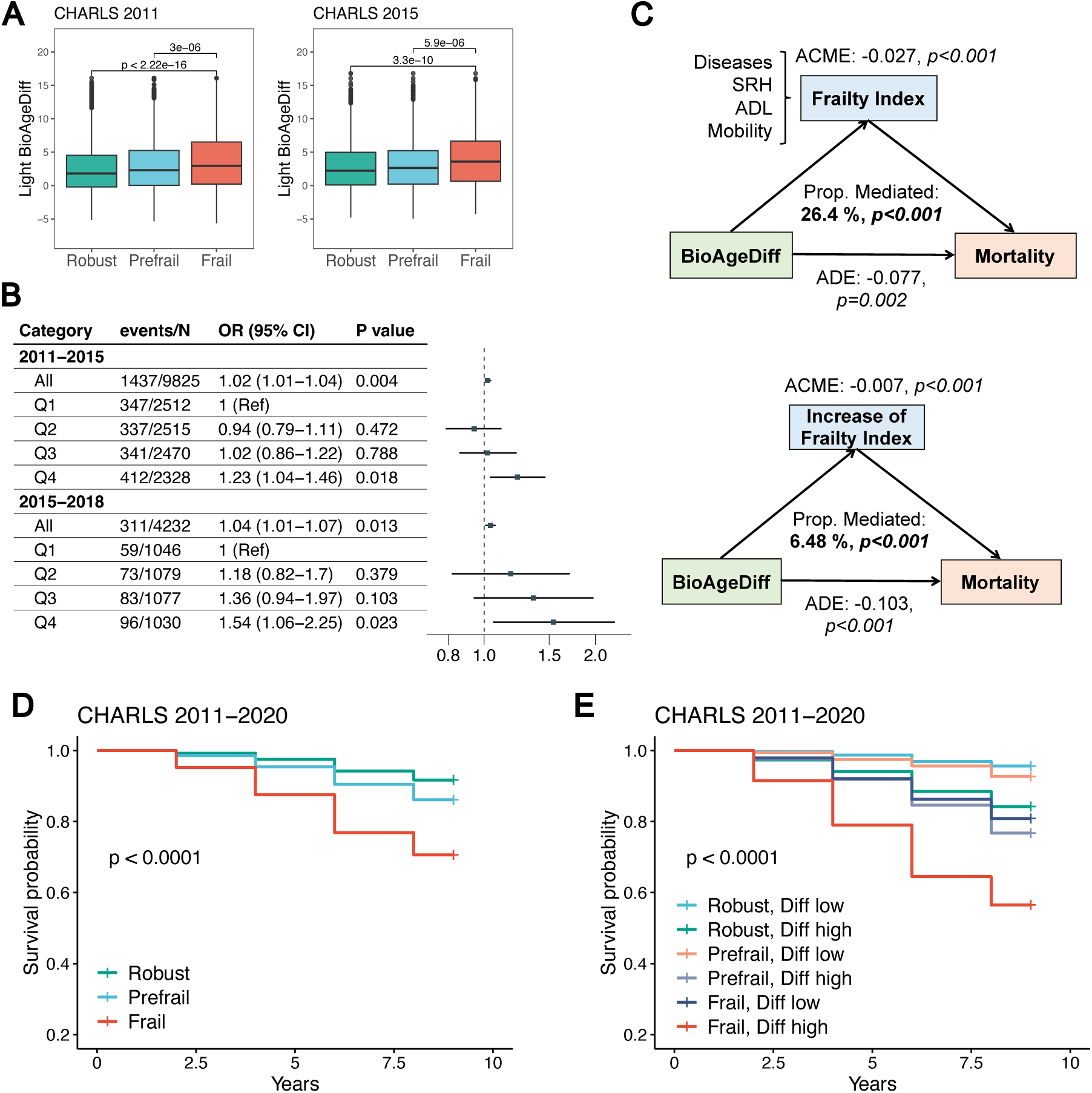
The Light BioAgeDiff, frailty and mortality in CHARLS. The boxplots (A) of Light BioAgeDiff across robust, prefrail, and frail individuals in CHARLS waves 1 (2011) and 3 (2015), with statistical significance determined using Wilcoxon tests. Forest plots (B) illustrated the associations between Light BioAgeDiff and incidence of frailty. The odd ratios were calculated through continuous and category Light BioAgeDiff (Q1-4: quantiles), adjusted by age and sex. The mediation models (C) of Light BioAge (wave 1, 2011), frailty index (wave 3, 2015) and mortality (wave 3-5, 2015-2022) were conducted. The change in frailty index was calculated based on assessments from waves 1 and 3. ADE: average direct effect; ACME: average causal mediated effect. The survival plots of individuals according to frailty status (D), and both frailty status and levels of Light BioAgeDiff (E).

The Light BioAgeDiff demonstrated performance comparable to that of frailty in predicting mortality, while the C-index of BioAgeDiff and frailty were 0.634 and 0.633 in CHARLS. using both BioAgeDiff and frailty as measures of biological and functional aging, we examined their combined effectiveness in identifying individuals at high risk for mortality. The survival probability of frail individuals was about 70% during 9-year follow-up in CHARLS (Figure 9D). In contrast, frail individuals with the highest BioAgeDiff had a mortality rate of approximately 55% during this period (Figure 9E). Therefore, These findings highlight the potential role of Light BioAgeDiff in preventing incident frailty and its joint contribution with frailty in identifying individuals at elevated risk for mortality.

## Discussion

In this study, we presented an elegant algorithm for estimating biological age as a linear combination of chronological age and various biomarkers. The GOLD biological age and its difference from chronological age provided insights into the relationship between individual biomarker values and the aging pace. Notably, the implementation of our algorithm in proteomics and metabolomics demonstrated the significant potential of omics biomarkers in identifying risks of mortality and and age-related chronic diseases. Furthermore, benchmark analysis demonstrated that our models outperformed traditional aging clocks in predicting the risks of both all-cause and cause-specific mortality across different age groups. We also developed a simplified version termed the Light BioAge, which provides a practical and efficient alternative with simplified calculations. The Light BioAge exhibited strong predictive capabilities in assessing mortality risks across three validation elderly cohorts and was associated with the onset of frailty, collectively forecasting mortality risks associated with frailty. In summary, our algorithm was validated as a general framework for constructing aging clocks. Importantly, both the BioAge and its light version can serve as convenient tools for aging assessment in clinical practice.

The robustness of the GOLD BioAge algorithm and the aging clocks were validated through multiple aspects. The evaluation of GOLD BioAge primarily focused on the correlation between BioAgeDiff and chronological age, the prediction of all-cause and cause-specific mortality, the incidence of multiple age-related chronic diseases, the onset of frailty, and validations across diverse populations. Additionally, benchmark analyses of mortality prediction demonstrated the superiority and sensitivity of the GOLD BioAge model. Consequently, GOLD BioAge served as a general and reliable measure of biological aging, offering simple and practical calculations for aging assessment and public health.

The pace of individual aging experiences dynamic changes throughout life, influenced by modifiable lifestyle choices, environmental factors, psychological influences, and health conditions. Identifying individuals at high risk of premature aging can enhance primary prevention efforts and reduce the healthcare and socioeconomic burdens linked to age-related diseases. In this study, the innovative biological aging clock exhibited stronger associations with morbidity and mortality than chronological age, providing a direct measure of an individual’s aging progression. To further advance the application of biological age in public health and clinical settings, we introduced the Light BioAge, a simple and practical aging clock that utilized just three accessible biomarkers alongside chronological age. The Light BioAge demonstrated applicability across various independent cohorts (NHANES, UKB, CHARLS, CLHLS, and RuLAS) with differing study designs, participant characteristics, and morbidity profiles. This model incorporated serum creatinine, blood glucose, and C-reactive protein levels with chronological age to reflect kidney function, metabolic and inflammatory status. These biomarkers were commonly used in medical examinations and were readily available at a low cost. Therefore, Light BioAge offered a convenient tool for ongoing monitoring of aging trajectories to prevent functional decline and age-related diseases.

Compared with the Levine’s phenotypic age, we both estimated the biological age by fitting the Gompertz distribution to empirical mortality data. The Levine’s phenotypic age had been widely used in aging-related studies. Notably, the phenotypic age outperformed earlier biological age-related methods in predicting all-cause mortality and various diseases^25^. GOLD BioAge exhibited a strong correlation with Levine’s phenotypic age in the NHANES and UKB dataset, demonstrating the robustness and reliability of our algorithm. Notably, the Levine’s phenotypic age relied on the Gompertz cumulative distribution function to estimate the 10-year mortality risk. Its calculation involved a double logarithmic transformation, which inevitably hindered its clinical interpretation. In comparison, our approach simplified the calculation process by utilizing the hazard function to identify instantaneous mortality risk, resulting in a higher predictive accuracy for mortality outcomes.

Our study introduced the ProtAge and MetAge, integrating omics data into the GOLD biological age framework. These novel aging clocks generally outperformed clinical marker-based clocks in predicting mortality, which probably due to the higher sensitivity of omics data in capture aging-related information^26^. For ProtAge, proteins were categorized into four groups based on their physiological function, each contributing to a distinct age estimate. Proteomics plays a crucial role in aging process, as changes in protein expression and post-translational modifications, particularly those linked to inflammation, oxidative stress, and cell cycle regulation, provide stable, long-term biomarkers. These molecular signatures offered deeper insights into biological aging compared to clinical markers. Additionally, because protein alterations often preceded the onset of chronic diseases, proteomics enhanced the early disease detection, making ProtAge a valuable tool for predicting mortality and early-stage health risks^27–29^. In addition, metabolomics reflected rapid, short-term fluctuations in the body’s biochemical processes, offering insights into how recent changes in diet, physical activity, and stress impact aging. The integration of both proteomic and metabolomics data into the aging clock framework created a more comprehensive tool to estimate biological age. It also provided the potential for personalized health interventions to mitigate aging-related risks.

This study has several limitations. First, although the omics-based aging clocks demonstrated superior performance compared to those using clinical biomarkers in the UKB dataset, further validation in other elderly cohorts is essential to confirm these findings. Additionally, the selection of biomarkers for the aging clocks was performed using LASSO penalized regression to enhance accuracy; however, different feature selection methods could yield alternative sets of biomarkers, indicating potential for further optimization of biomarker panels in clinical applications. Furthermore, we validated the Light BioAge in three Chinese cohorts, leaving uncertain whether the full GOLD BioAge model would more accurately capture the risks associated with geriatric syndromes and mortality.

## Methods and materials

### Study populations

Our study used the data of NHANES 1999-2018, UKB, CHARLS, CLHLS, and RuLAS. The US NHANES is a nationally representative cross-sectional survey of civilian living in the US, approved by the National Center for Health Statistics (NCHS) Ethics Review Board^30^. The UK Biobank is large-scale perspective cohort that collected data from over 500,000 participants across 22 centers in England, Scotland, and Wales. UKB received ethics approval from the North West Multicenter Research Ethics Commitee^31^. The CHARLS is an ongoing prospective population-based longitudinal cohort study of middle-aged and older Chinese adults. CHARLS was approved by the Ethics Review Board of Peking University, which was conducted in accordance with the Declaration of Helsinki and other relevant guidelines and regulations^32^. The CLHLS is a nationwide longitudinal study of old-aged Chinese population. The project was approved by the Biomedical Ethics Committee of Peking University, China (IRB00001052-13074)^33^. The Rugao Longevity and Ageing Study (RuLAS) is a population-based perspective study, which consisted of a longevity cohort and an aging cohort in Rugao, China^34^. The RuLAS was approved by the Human Ethics Committee of Fudan University School of Life Sciences. All participants provided written informed consent. And This study followed the Strengthening the Reporting of Observational Studies in Epidemiology (STROBE) reporting guidelines for cohort studies^35^.

### Clinical biomarker selection for constructing Gold BioAge

We utilized the data of NHANES 1999-2018 for variable selection, and refined the biomarker panel for constructing Gold Biological Age. As a result, 26 common biomarkers from cell blood count (CBC) tests and biochemical assays were enrolled (**Table S1**). LASSO Cox regression models were employed for biomarker selection, with five-fold cross-validation to determine the optimal parameter value (lambda.1se) of 0.0166. First, the 26 biomarkers and chronological age were analyzed using the LASSO Cox regression. Then 10 biomarkers were retained in the model (**Figure S1**), including chronological age, creatinine, glucose, among others. This set of biomarkers formed the basis for the novel biological age model (Gold BioAge). To simplify the panel for practical use, feature selection was performed on 10 blood biomarkers (biochemical and hematological) that were consistently observed across various cohorts (**Table S2**). This simplified panel included chronological age, serum creatinine, glucose, and C-reactive protein (CRP) for the light Gold Biological age model (Light BioAge).

### Metabolomics and proteomics biomarker selection

Then, we applied GOLD BioAge models to metabolic and proteomic biomarkers, respectively, employing data from the UK Biobank (UKB, 2006-2010). For the Gold Proteomic Age Model (Gold ProtAge), we analyzed 2,923 proteins from 53,014 participants. To maintain the relative integrity of the independent sample, we removed proteins with more than 10% missing data, resulting in 1,459 protein biomarkers for analysis. Using LASSO-Cox regression model, we calculated protein-predicted age (ProtAge) in the entire sample (n = 39,772) through five-fold cross-validation. To achieve a balance between simplicity and predictive power, we opted for a lambda value of exp (−6), optimizing the model’s simplicity and model performance (**Figure S1**). Then it selected 22 protein biomarkers along with chronological age. Detailed descriptions of all the selected proteins are available in the supplementary materials. For the Gold Metabolic Aging Clock (Gold MetAge), we utilized nuclear magnetic resonance (NWR) - based blood profiling metabolomics data from UKB. A total of 248,202 UKB participants were enrolled, each with measurements of 251 circulating metabolomic markers. We performed LASSO-Cox regression using fivefold cross-validation, selecting features based on the lambda value of exp (−6), which corresponded to a one standard deviation increase over the lambda with minimum mean-squared error. Among the 251 metabolic biomarkers, 27 were selected, along with chronological age, to develop the Gold MetAge.

### GOLD BioAge model training

We conducted two Gompertz regression models for biological age model training. The first Gompertz regression model only included chronological age as a predictor of time-to-mortality data. The second Gompertz regression model incorporated both chronological age and selected biomarkers as predictors. We defined the gold biological age (Gold BioAge) as the age accounting for the actual mortality hazard by considering both chronological age (CA) and additional biomarkers (**Figure S2**). The models were specified as follows:

Model 1: Chronological Age Only:

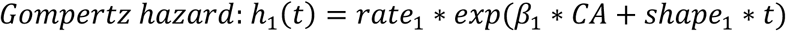

Model 2: Chronological Age and Selected Biomarkers:

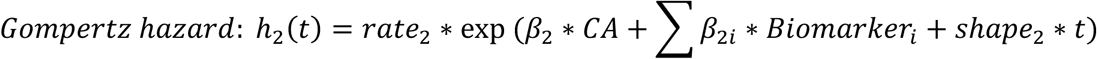

In Model 2, biomarkers represented the selected biomarkers included in the model, and β_i_ (coef) represented the coefficients of each biomarker. The GOLD BioAge integrated chronological age with relevant biomarkers to better reflect mortality hazard and aging status. Let *h*_1_(*Bioage*, *t* = 0) ≈ *h*_2_(*Bioage*, *Biomarkers*, *t* = 0). In real dataset, the *h*_1_ and *h*_2_ empirical distributions were different (Figure S2), resulting in underestimate of biological age in the whole population. Thus, we add a constant (γ) correct the bias and let *h*_1_ = γ ∗ *h*_2_.

Thus, Gold BioAge was derived as follows:

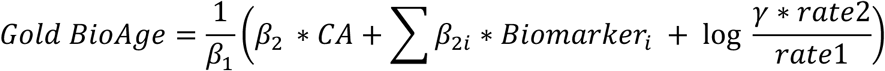

For further simplicity and robustness, we set the coefficient parameter of CA (β_2_) equal to the parameter (β_1_) in Model 2, and estimated the parameter for rate, shape, and coefficients of biomarkers.

When β_1_ = β_2_, the formula was further simplified as follows:

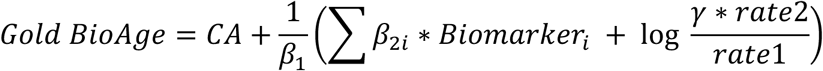

The Gompertz distribution parameters (rate, shape, and coefficients) were estimated by maximum likelihood using the “flexsurv” R package. The coefficients of GOLD BioAge, ProtAge and MetAge were shown in the **Table S5-8**.The algorithm of GOLD BioAge was implemented as a R package (http://github.com/Jerryhaom/GOLDBioAge).

### Benchmark of biological age models

To ensure the robustness of our models, we evaluated the model performance in the NHANES and UKB, compared with other common phenotypic aging clocks and dimensional reduction methods. The the Levine phenotypic age, KDM age and mahalanobis distance statistic were calculated using the ‘BioAge’ R package^36^. The PCA age was calculated through principal component analysis and regressed to age with first five components. In the NHANES and UKB datasets, we compared the concordance index (C-index) of survival analysis and Area Under the Curve (AUC) of 10-year mortality prediction in the full samples and across three age groups: young adults, middle-aged adults, and older adults. It allowed us to determine the models’ ability to discriminate mortality risk across different age groups. In addition to all-cause mortality, these aging clocks in predicting cause-specific mortality were evaluated. Since the aging clock was a single variable, the prediction values of 10-year survival status was estimated through on logistic regressions based on aging clcoks. This comprehensive benchmarking analysis allowed for a thorough evaluation of the models’ performance and comparison with other established aging clocks. The associations of GOLD BioAge, Light BioAge, ProtAge, and MetAge with all-cause and cause-specific mortality were shown in **Table S9-11**.

### Validation in independent elderly cohorts

We validated the Light BioAge model in other three elderly cohorts: the CHARLS, RuLAS and CLHLS datasets. Five waves of CHARLS data (2011-2020) were utilized, with blood-based bioassay data used to construct the Light BioAge. Health and function questionnaires were collected for frailty assessment^37^ (**Table S12**). For RuLAS, the wave 2 (2016) was taken as baseline, and data of blood biomarkers were obtained. The data CLHLS 2014-2018 was enrolled to validate the LightBioAge. We calculated the ROC curves to evaluate the prediction performance of Light BioAge and chronological age across the all sample, as well as subpopulations stratified by age (60-79 years; >= 80 years). Additionally, we fitted survival curves for low-risk and high-risk groups based on the Light BioAgeDiff model.

### Assessment of mortality and onset of chronic diseases

In NHANES, death information was based on linked data from records taken from the National Death Index (NDI) through December 31, 2019, provided through the Centers for Disease Control and Prevention. Data on mortality status and length of follow-up (in person-months) were available for nearly all participants. In UKB, death information was obtained through death certificates held within the National Health Service (NHS) Information Centre (England and Wales) and the NHS Central Register (Scotland) to November 30, 2022. We calculated participants’ time to death from baseline to the date of death, date of loss to follow-up, or date of last record of follow-up, whichever came first. We used the International Statistical Classification of Diseases, 10th, to define causes of death. The cause-specific mortality included mortality of malignant neoplasm, heart disease, cerebrovascular disease, respiratory disease, Alzheimer disease, diabetes, and others.

In addition, diagnosed dates of incident chronic disease in UKB were also collected, including cancer, myocardial infraction, heart failure, stroke, chronic obstructive pulmonary disease (COPD), and dementia.

### Assessment of health-related factors and outcomes

The unhealthy lifestyle score was based on six modifiable lifestyle factors: smoking, alcohol consumption, physical activity, diet, body mass index (BMI), and sedentary behavior, defined by World Health Organization.The score was categorized into five groups (0, 1, 2, 3, 4 and more unhealthy factors). Multimobidites, defined as the number of lifetime disease diagnoses. In NHANES, we included diabetes, high blood pressure, congestive heart failure, coronary heart disease, heart attack, stroke, cancer or malignancy, and chronic bronchitis; In UKB, we included cancer, myocardial infarction, heart failure, stroke, chronic obstructive pulmonary disease (COPD), and dementia. Disease count was classified into five categories: no disease, 1, 2, 3, and 4 or more diseases. Self-rated health was recorded in four levels: excellent or very good, good, fair and poor. The distributions of GOLD BioAge, Light BioAge, ProtAge, and MetAge by unhealthy lifestyles, comorbidity, and self-rated health were shown in **Table S13**.

### Statistical analysis

Survival analysis was conducted in different age groups. Within the same group, participants were classified into quintiles based on their BioAge Difference (BioAgeDiff), with the top 20% representing individuals at highest risk of death. Kaplan-Meier survival curves were then plotted to compare the predicted survival probabilities between the highest and lowest quintiles of the novel BioAge. Harrell’s Concordance Index (C-index) was used to assess the predictive discrimination in survival analysis. And the Area Under the Curve (AUC) was took as a robust metric to evaluate the prediction ability. Cox proportional hazard models were conducted to assess the associations between different biological aging clocks, mortality and the onset of chronic diseases. The cox models were adjusted for sex and chronological age. All statistical analyses were performed using R version 4.3.3.

## Data Availability

The data of RulAS are available through reasonable request from the corresponding author. The data from CLHLS are available at https://opendata.pku.edu.cn/dataset.xhtml. The data from CHARLS are available at https://charls.charlsdata.com/pages/data/111/zh-cn.html. The data from the NHANES are available at www.cdc.gov/nchs/nhis/index.htm, and the data from the UK Biobank are available upon application at www.ukbiobank.ac.uk/register-apply. This research was conducted using UK Biobank Resource under Application Number 103791.

## Acknowledgments

The data used in this research were obtained from the NHANES, UK Biobank, CHARLS, RuLAS and CLHLS. We would like to thank the workers, researchers, and participants involved in these cohorts.

## Author contributions

Concept and design: Meng Hao, Li Yi, Hui Zhang.

Acquisition, analysis, or interpretation of data: Meng Hao, Zixin Hu, Shuai Jiang. Drafting of the manuscript: Meng Hao, Jingyi Wu, Hui Zhang

Critical revision of the manuscript for important intellectual content: All authors. Statistical analysis: Meng Hao, Hui Zhang, Jingyi Wu.

Administrative, technical, or material support: Xiangnan Li, Shuming Wang, Meijia Wang, Yaqi Huang, Jiaofeng Wang, Jie Chen, Zhijun Bao, Li Jin.

Supervision: Meng Hao, Yi Li, Shuai Jiang, Zixin Hu, Xiaofeng Wang.

## References

1 Ferrucci, L., Levine, M. E., Kuo, P. L. & Simonsick, E. M. Time and the Metrics of Aging. Circ Res 123, 740–744 (2018). 10.1161/CIRCRESAHA.118.312816

2 Ferrucci, L. & Kuchel, G. A. Heterogeneity of Aging: Individual Risk Factors, Mechanisms, Patient Priorities, and Outcomes. J Am Geriatr Soc 69, 610–612 (2021). 10.1111/jgs.17011

3 Moqri, M. et al. Validation of biomarkers of aging. Nat Med 30, 360–372 (2024). 10.1038/s41591-023-02784-9

4 Guo, J. et al. Aging and aging-related diseases: from molecular mechanisms to interventions and treatments. Signal Transduct Target Ther 7, 391 (2022). 10.1038/s41392-022-01251-0

5 Hartmann, A. et al. Ranking Biomarkers of Aging by Citation Profiling and Effort Scoring. Front Genet 12, 686320 (2021). 10.3389/fgene.2021.686320

6 Jylhävä, J., Pedersen, N. L. & Hägg, S. Biological Age Predictors. eBioMedicine 21, 29–36 (2017). 10.1016/j.ebiom.2017.03.046

7 Moqri, M. et al. Biomarkers of aging for the identification and evaluation of longevity interventions. Cell 186, 3758–3775 (2023). 10.1016/j.cell.2023.08.003

8 Levine, M. E. et al. An epigenetic biomarker of aging for lifespan and healthspan. Aging (Albany NY) 10, 573–591 (2018). 10.18632/aging.101414

9 Fong, S. et al. Principal component-based clinical aging clocks identify signatures of healthy aging and targets for clinical intervention. Nat Aging 4, 1137–1152 (2024). 10.1038/s43587-024-00646-8

10 Qiu, W., Chen, H., Kaeberlein, M. & Lee, S. I. ExplaiNAble BioLogical Age (ENABL Age): an artificial intelligence framework for interpretable biological age. Lancet Healthy Longev 4, e711–e723 (2023). 10.1016/S2666-7568(23)00189-7

11 Rutledge, J., Oh, H. & Wyss-Coray, T. Measuring biological age using omics data. Nature Reviews Genetics 23, 715–727 (2022). 10.1038/s41576-022-00511-7

12 Horvath, S. & Raj, K. DNA methylation-based biomarkers and the epigenetic clock theory of ageing. Nature reviews genetics 19, 371–384 (2018).

13 McCrory, C. et al. GrimAge Outperforms Other Epigenetic Clocks in the Prediction of Age-Related Clinical Phenotypes and All-Cause Mortality. The Journals of Gerontology: Series A 76, 741–749 (2020). 10.1093/gerona/glaa286

14 Oh, H. S. et al. Organ aging signatures in the plasma proteome track health and disease. Nature 624, 164–172 (2023). 10.1038/s41586-023-06802-1

15 Tian, Y. E. et al. Heterogeneous aging across multiple organ systems and prediction of chronic disease and mortality. Nature Medicine 29, 1221–1231 (2023). 10.1038/s41591-023-02296-6

16 Argentieri, M. A. et al. Proteomic aging clock predicts mortality and risk of common age-related diseases in diverse populations. Nat Med 30, 2450–2460 (2024). 10.1038/s41591-024-03164-7

17 Jia, X. et al. A Novel Metabolomic Aging Clock Predicting Health Outcomes and Its Genetic and Modifiable Factors. Adv Sci (Weinh), e2406670 (2024). 10.1002/advs.202406670

18 Zhang, S. et al. A metabolomic profile of biological aging in 250,341 individuals from the UK Biobank. Nat Commun 15, 8081 (2024). 10.1038/s41467-024-52310-9

19 Kuo, C. L. et al. Proteomic aging clock (PAC) predicts age-related outcomes in middle-aged and older adults. Aging Cell 23, e14195 (2024). 10.1111/acel.14195

20 Biomarkers of Aging, C., et al. Challenges and recommendations for the translation of biomarkers of aging. Nat Aging (2024). 10.1038/s43587-024-00683-3

21 Gompertz, B. XXXIII. A supplement to two papers published in the Transactions of the Royal Society, “On the science connected with human mortality;” the one published in 1820, and the other in 1825. Philosophical Transactions of the Royal Society of London 152, 511–559 (1862). doi:10.1098/rstl.1862.0026

22 Cohen, A. A. et al. A novel statistical approach shows evidence for multi-system physiological dysregulation during aging. Mech Ageing Dev 134, 110–117 (2013). 10.1016/j.mad.2013.01.004

23 Li, Q. et al. Homeostatic dysregulation proceeds in parallel in multiple physiological systems. Aging Cell 14, 1103–1112 (2015). 10.1111/acel.12402

24 Fong, S. et al. Principal component-based clinical aging clocks identify signatures of healthy aging and targets for clinical intervention. Nature Aging, 1–16 (2024).

25 Liu, Z. et al. A new aging measure captures morbidity and mortality risk across diverse subpopulations from NHANES IV: A cohort study. PLoS Med 15, e1002718 (2018). 10.1371/journal.pmed.1002718

26 Argentieri, M. A. et al. Proteomic aging clock predicts mortality and risk of common age-related diseases in diverse populations. Nature Medicine 30, 2450–2460 (2024). 10.1038/s41591-024-03164-7

27 Kuo, C. L. et al. Proteomic aging clock (PAC) predicts age-related outcomes in middle-aged and older adults. Aging Cell 23, e14195 (2024).

28 Perry, A. S. et al. Proteomic analysis of cardiorespiratory fitness for prediction of mortality and multisystem disease risks. Nature Medicine, 1–11 (2024).

29 Carrasco-Zanini, J. et al. Proteomic signatures improve risk prediction for common and rare diseases. Nature medicine, 1–10 (2024).

30 Nguyen, V. K. et al. Harmonized US National Health and Nutrition Examination Survey 1988-2018 for high throughput exposome-health discovery. medRxiv (2023). 10.1101/2023.02.06.23284573

31 Sudlow, C. et al. UK biobank: an open access resource for identifying the causes of a wide range of complex diseases of middle and old age. PLoS medicine 12, e1001779 (2015).

32 Zhao, Y., Hu, Y., Smith, J. P., Strauss, J. & Yang, G. Cohort profile: the China health and retirement longitudinal study (CHARLS). International journal of epidemiology 43, 61–68 (2014).

33 Gu, D., Feng, Q., Chen, H. & Zeng, Y. in Encyclopedia of gerontology and population aging 957-970 (Springer, 2022).

34 Hao, M. et al. Phenotype correlations reveal the relationships of physiological systems underlying human ageing. Aging Cell 20, e13519 (2021). 10.1111/acel.13519

35 Liu, Z. et al. Cohort profile: the Rugao longevity and ageing study (RuLAS). International journal of epidemiology 45, 1064–1073 (2016).

36 Kwon, D. & Belsky, D. W. A toolkit for quantification of biological age from blood chemistry and organ function test data: BioAge. Geroscience 43, 2795–2808 (2021). 10.1007/s11357-021-00480-5

37 He, D. et al. Changes in frailty and incident cardiovascular disease in three prospective cohorts. Eur Heart J 45, 1058–1068 (2024). 10.1093/eurheartj/ehad885

## References

1 Leon-Gonzalez, R. et al. Growth Differentiation Factor 15 as a Biomarker of Cardiovascular Risk in Chronic Musculoskeletal Pain. J Gerontol A Biol Sci Med Sci 79 (2024). 10.1093/gerona/glae163

2 Li, J. et al. Overview of growth differentiation factor 15 (GDF15) in metabolic diseases. Biomed Pharmacother 176, 116809 (2024). 10.1016/j.biopha.2024.116809

3 Ababneh, O., Nishizaki, D., Kato, S. & Kurzrock, R. Tumor necrosis factor superfamily signaling: life and death in cancer. Cancer Metastasis Rev (2024). 10.1007/s10555-024-10206-6

4 Dostert, C., Grusdat, M., Letellier, E. & Brenner, D. The TNF Family of Ligands and Receptors: Communication Modules in the Immune System and Beyond. Physiol Rev 99, 115–160 (2019). 10.1152/physrev.00045.2017

5 Chen, Y. et al. WAP four-disulfide core domain protein 2 mediates the proliferation of human ovarian cancer cells through the regulation of growth- and apoptosis-associated genes. Oncol Rep 29, 288–296 (2013). 10.3892/or.2012.2114

6 Zhao, D., Song, J. & Ji, C. Endoplasmic reticulum stress regulates apoptosis and chemotherapeutic via enhancing TNFRSF10B recycling to the cell membrane in triple-negative breast cancer. Clin Transl Oncol (2024). 10.1007/s12094-024-03509-1

7 Arosa, L. et al. RNA Expression of MMP12 Is Strongly Associated with Inflammatory Bowel Disease and Is Regulated by Metabolic Pathways in RAW 264.7 Macrophages. Int J Mol Sci 25 (2024). 10.3390/ijms25063167

8 Kuntschar, S. et al. Mmp12 Is Translationally Regulated in Macrophages during the Course of Inflammation. Int J Mol Sci 24 (2023). 10.3390/ijms242316981

9 Kalatha, T. et al. Does cognitive dysfunction correlate with neurofilament light polypeptide levels in the CSF of patients with multiple sclerosis? J Int Med Res 47, 2187–2198 (2019). 10.1177/0300060519840550

10 Zhao, Y., Arceneaux, L., Culicchia, F. & Lukiw, W. J. Neurofilament Light (NF-L) Chain Protein from a Highly Polymerized Structural Component of the Neuronal Cytoskeleton to a Neurodegenerative Disease Biomarker in the Periphery. HSOA J Alzheimers Neurodegener Dis 7 (2021). 10.24966/AND-9608/100056

11 Zhao, N. et al. CUB Domain-Containing Protein 1 (CDCP1) Is a Target for Radioligand Therapy in Castration-Resistant Prostate Cancer, including PSMA Null Disease. Clin Cancer Res 28, 3066–3075 (2022). 10.1158/1078-0432.CCR-21-3858

12 Lim, S. A. et al. Targeting a proteolytic neoepitope on CUB domain containing protein 1 (CDCP1) for RAS-driven cancers. J Clin Invest 132 (2022). 10.1172/JCI154604

13 Zhang, D., Lu, H., Hou, W., Bai, Y. & Wu, X. Effect of miR-132-3p on sepsis-induced acute kidney injury in mice via regulating HAVCR1/KIM-1. Am J Transl Res 13, 7794–7803 (2021).

14 Song, J. et al. Understanding kidney injury molecule 1: a novel immune factor in kidney pathophysiology. Am J Transl Res 11, 1219–1229 (2019).

15 Aggarwal, S., Dabla, P. K. & Arora, S. Prostasin: An Epithelial Sodium Channel Regulator. J Biomark 2013, 179864 (2013). 10.1155/2013/179864

16 Rickert, K. W. et al. Structure of human prostasin, a target for the regulation of hypertension. J Biol Chem 283, 34864–34872 (2008). 10.1074/jbc.M805262200

17 Shi, Y. et al. Latent-transforming growth factor beta-binding protein-2 (LTBP-2) is required for longevity but not for development of zonular fibers. Matrix Biol 95, 15–31 (2021). 10.1016/j.matbio.2020.10.002

18 Zou, M. et al. Plasma LTBP2 as a potential biomarker in differential diagnosis of connective tissue disease-associated interstitial lung disease and idiopathic pulmonary fibrosis: a pilot study. Clin Exp Med 23, 4809–4816 (2023). 10.1007/s10238-023-01214-x

19 Sheehan, S. M. & Allen, R. E. Skeletal muscle satellite cell proliferation in response to members of the fibroblast growth factor family and hepatocyte growth factor. J Cell Physiol 181, 499–506 (1999). 10.1002/(SICI)1097-4652(199912)181:3<499::AID-JCP14>3.0.CO;2-1

20 Wilson, S. E. et al. Effect of epidermal growth factor, hepatocyte growth factor, and keratinocyte growth factor, on proliferation, motility and differentiation of human corneal epithelial cells. Exp Eye Res 59, 665–678 (1994). 10.1006/exer.1994.1152

21 Matsumoto, K., Umitsu, M., De Silva, D. M., Roy, A. & Bottaro, D. P. Hepatocyte growth factor/MET in cancer progression and biomarker discovery. Cancer Sci 108, 296–307 (2017). 10.1111/cas.13156

22 Kishore, A., Purcell, R. H., Nassiri-Toosi, Z. & Hall, R. A. Stalk-dependent and Stalk-independent Signaling by the Adhesion G Protein-coupled Receptors GPR56 (ADGRG1) and BAI1 (ADGRB1). J Biol Chem 291, 3385–3394 (2016). 10.1074/jbc.M115.689349

23 Damkjaer, M. et al. Renal renin secretion as regulator of body fluid homeostasis. Pflugers Arch 465, 153–165 (2013). 10.1007/s00424-012-1171-2

24 Schmieder, R. E., Hilgers, K. F., Schlaich, M. P. & Schmidt, B. M. Renin-angiotensin system and cardiovascular risk. Lancet 369, 1208–1219 (2007). 10.1016/S0140-6736(07)60242-6

25 McKie, P. M. & Burnett, J. C., Jr. NT-proBNP: The Gold Standard Biomarker in Heart Failure. J Am Coll Cardiol 68, 2437–2439 (2016). 10.1016/j.jacc.2016.10.001

26 Booth, R. A. et al. Performance of BNP and NT-proBNP for diagnosis of heart failure in primary care patients: a systematic review. Heart Fail Rev 19, 439–451 (2014). 10.1007/s10741-014-9445-8

27 Wee, P. & Wang, Z. Epidermal Growth Factor Receptor Cell Proliferation Signaling Pathways. Cancers (Basel) 9 (2017). 10.3390/cancers9050052

28 Zhao, W. et al. Epidermal growth factor receptor mutations and brain metastases in non-small cell lung cancer. Front Oncol 12, 912505 (2022). 10.3389/fonc.2022.912505

29 Chan, C. M. et al. Carcinoembryonic Antigen-Related Cell Adhesion Molecule 5 Is an Important Surface Attachment Factor That Facilitates Entry of Middle East Respiratory Syndrome Coronavirus. J Virol 90, 9114–9127 (2016). 10.1128/JVI.01133-16

30 Frischknecht, R. & Seidenbecher, C. I. Brevican: a key proteoglycan in the perisynaptic extracellular matrix of the brain. Int J Biochem Cell Biol 44, 1051–1054 (2012). 10.1016/j.biocel.2012.03.022

31 Carim-Todd, L., Escarceller, M., Estivill, X. & Sumoy, L. LRRN6A/LERN1 (leucine-rich repeat neuronal protein 1), a novel gene with enriched expression in limbic system and neocortex. Eur J Neurosci 18, 3167–3182 (2003). 10.1111/j.1460-9568.2003.03003.x

32 Liu, B. et al. Leucine-rich repeat neuronal protein-1 suppresses apoptosis of gastric cancer cells through regulation of Fas/FasL. Cancer Sci 110, 2145–2155 (2019). 10.1111/cas.14042

33 Peeters, M. C. et al. The adhesion G protein-coupled receptor G2 (ADGRG2/GPR64) constitutively activates SRE and NFkappaB and is involved in cell adhesion and migration. Cell Signal 27, 2579–2588 (2015). 10.1016/j.cellsig.2015.08.015

34 Corda, P. O., Santiago, J. & Fardilha, M. G-Protein Coupled Receptors in Human Sperm: An In Silico Approach to Identify Potential Modulatory Targets. Molecules 27 (2022). 10.3390/molecules27196503

35 Gevaert, T. et al. The stem cell growth factor receptor KIT is not expressed on interstitial cells in bladder. J Cell Mol Med 21, 1206–1216 (2017). 10.1111/jcmm.13054

36 Miettinen, M., Kraszewska, E., Sobin, L. H. & Lasota, J. A nonrandom association between gastrointestinal stromal tumors and myeloid leukemia. Cancer 112, 645–649 (2008). 10.1002/cncr.23216

37 Lin, Y. et al. Serum insulin-like growth factor-I, insulin-like growth factor binding protein-3, and the risk of pancreatic cancer death. Int J Cancer 110, 584–588 (2004). 10.1002/ijc.20147

38 Ruan, W. & Lai, M. Insulin-like growth factor binding protein: a possible marker for the metabolic syndrome? Acta Diabetol 47, 5–14 (2010). 10.1007/s00592-009-0142-3

39 Imani, F. et al. Advanced glycosylation endproduct-specific receptors on human and rat T-lymphocytes mediate synthesis of interferon gamma: role in tissue remodeling. J Exp Med 178, 2165–2172 (1993). 10.1084/jem.178.6.2165

40 Prasad, K. & Mishra, M. AGE-RAGE Stress, Stressors, and Antistressors in Health and Disease. Int J Angiol 27, 1–12 (2018). 10.1055/s-0037-1613678

41 Goulart, A. C., Germer, S., Rexrode, K. M., Martin, M. & Zee, R. Y. Polymorphisms in advanced glycosylation end product-specific receptor (AGER) gene, insulin resistance, and type 2 diabetes mellitus. Clin Chim Acta 398, 95–98 (2008). 10.1016/j.cca.2008.08.020

