## Supplementary figures for "Gompertz law based biological age (GOLD BioAge): a simple and practical measurement of biological aging to capture morbidity and mortality risks"

**Figure S1.** LASSO Cox regression penalty in selecting features to develop GOLD aging clocks.

**Figure S2.** Estimate of GOLD BioAge and BioAgeDiff through Gompertz mortality hazard.

**Figure S3.** Validation of GOLD BioAge in UKB.

**Figure S4.** GOLD ProtAgeDiff and MetAgeDiff in UKB.

**Figure S5.** Associations of ProtAgeDiff, MetAgeDiff, and BioAgeDiff with mortality in UKB.

**Figure S6.** The predictive capability of ProtAge and its subpanels regarding mortality.

**Figure S7.** The comparison of GOLD BioAge, and other common aging clocks in predicting mortality in NHANES III.

**Figure S8.** The mortality hazard, Light BioAge and Levine's phenotypic age in NHANES.

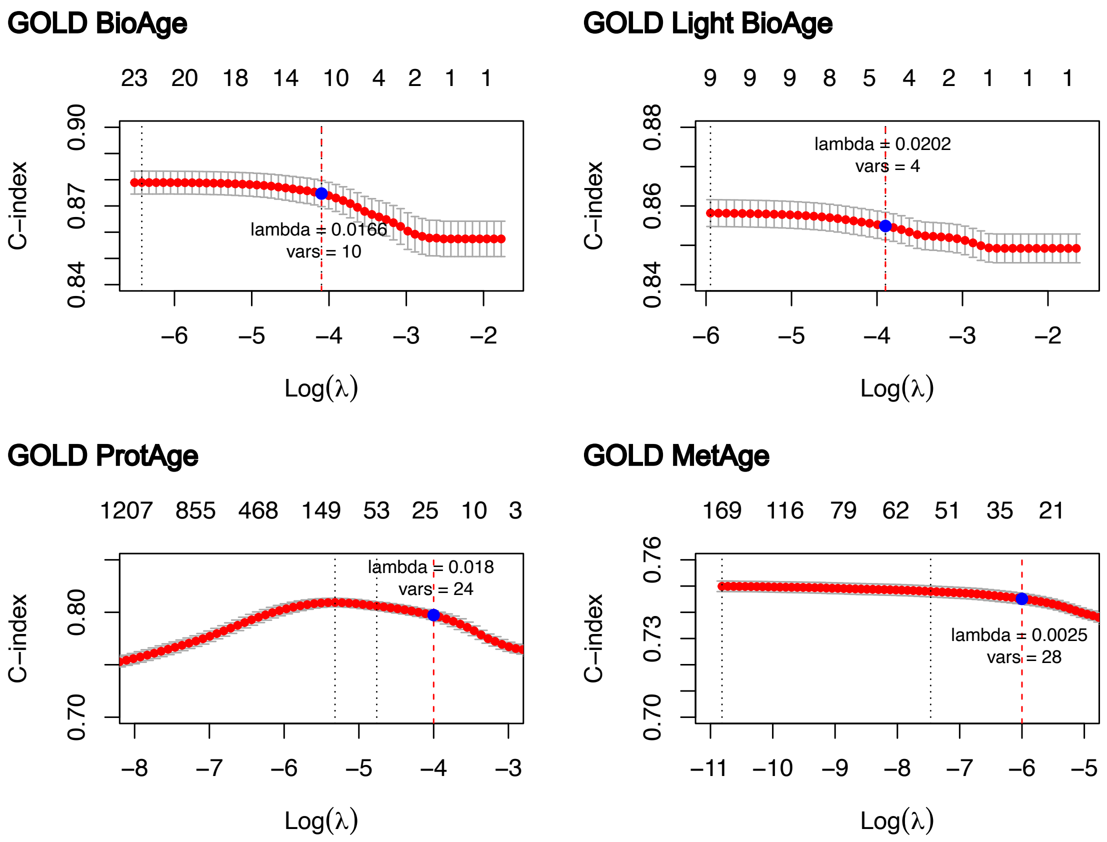


**Figure S1. LASSO Cox regression penalty in selecting features to develop GOLD aging clocks.** Harrel's concordance measure (C-index) for mortality was assessed across various levels of penalty (lambda, λ). The lambda value was chosen to be as high as possible to select fewer features while maintaining a relatively high C-index. Optimal lambda.1se values and higher values were determined for each aging clocks: BioAge with λ = 0.0166 and 10 variables selected; Light BioAge with λ = 0.0202 and 4 variables selected; ProtAge with λ = 0.018 and 24 variables selected; MetAge with λ = 0.0025 and 28 variables selected.

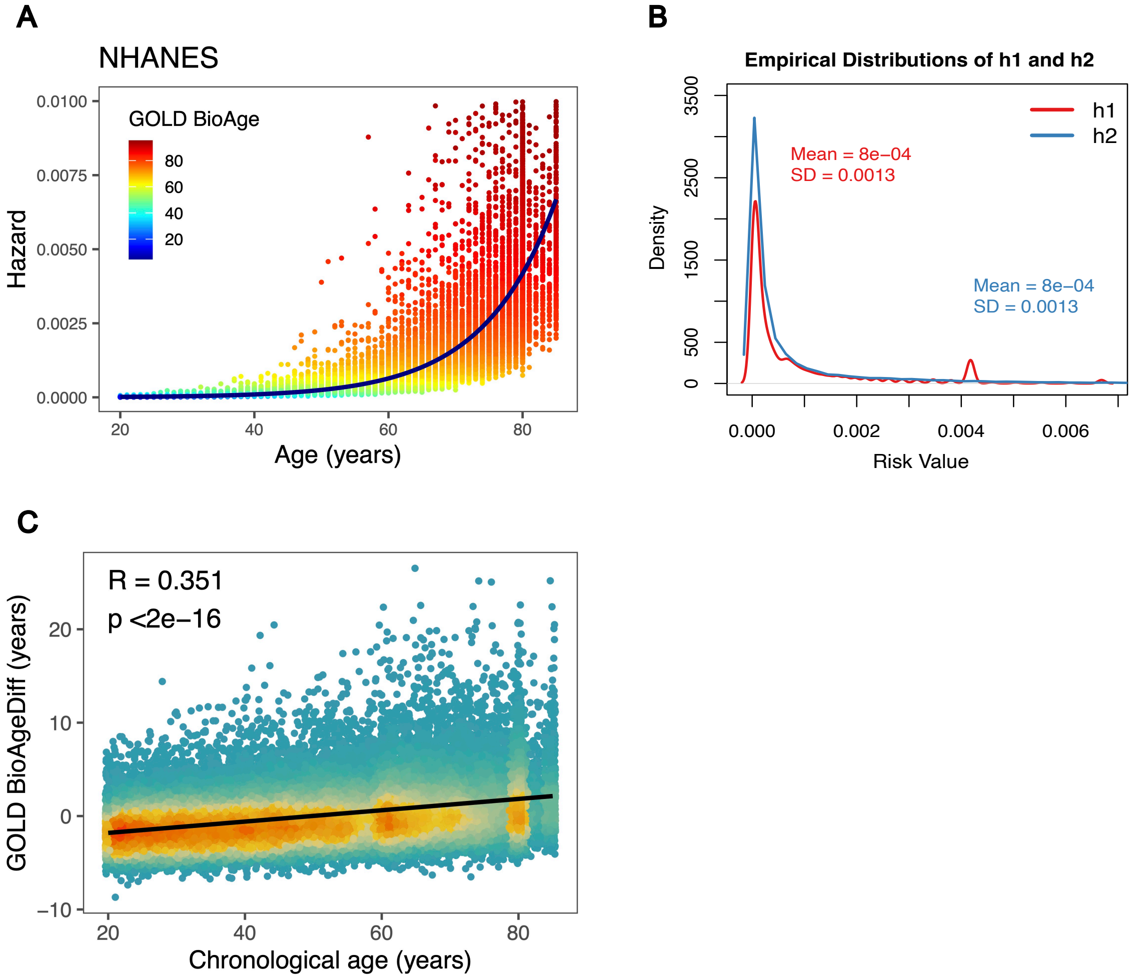


**Figure S2. Estimate of GOLD BioAge and BioAgeDiff through Gompertz mortality hazard.** Two Gompertz regression models (Methods, Model 1, Model 2) were used to predict mortality hazards for each individual in NHANES (A). The mortality hazards, considering both chronological age and biomarkers, were depicted as colorful points (Model 2), while the navy line represented the exponential increase of mortality hazard with chronological age (Model 1). GOLD BioAge was defined as the age corresponding to the mortality hazard of the points. The empirical distribution of mortality hazard function of the two models were shown (B). GOLD BioAgeDiff was calculated as the differences between the mortality hazard values of the line and points, showing a positive correlation with age (B).

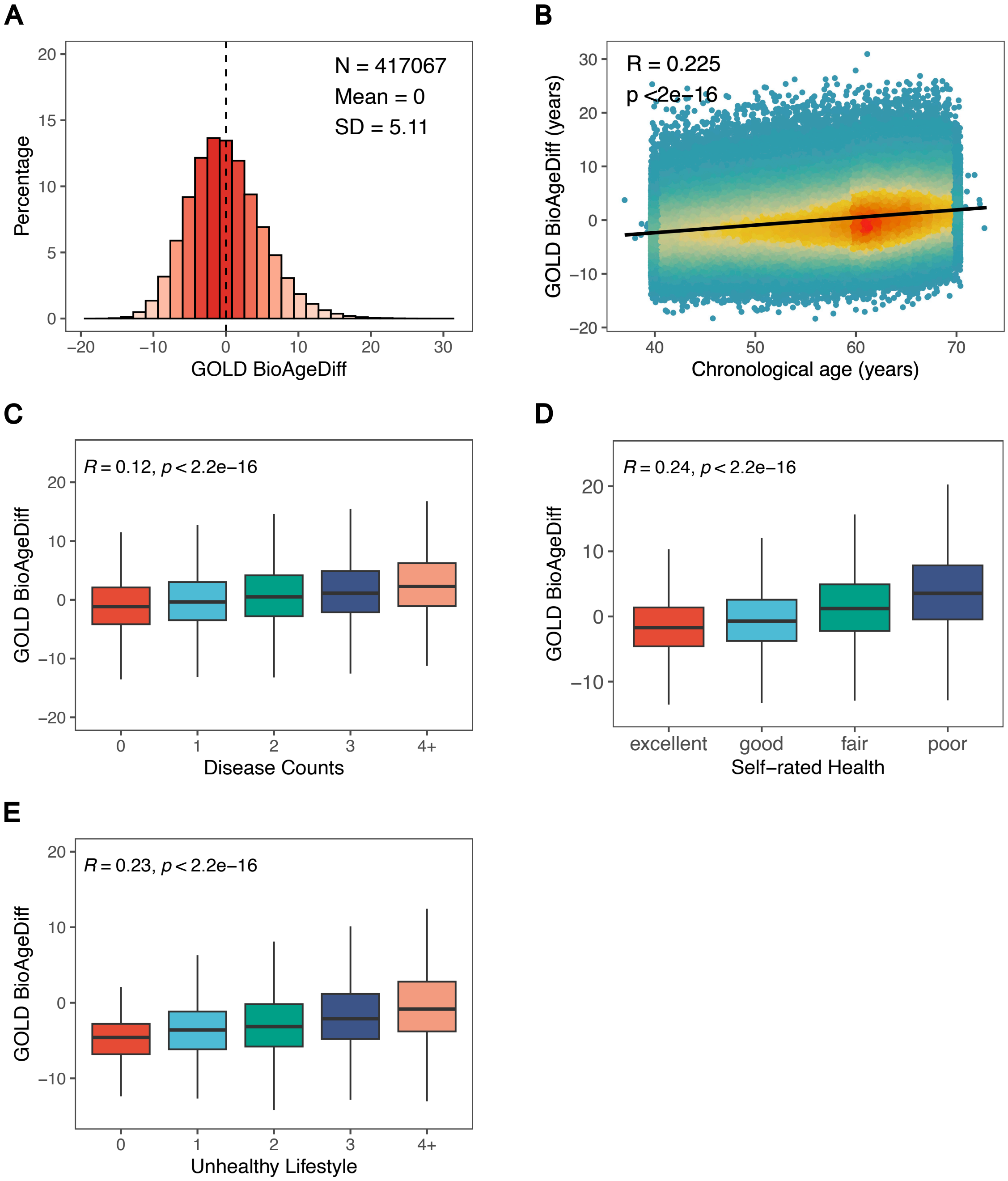


**Figure S3. Validation of GOLD BioAge in UKB.** The histogram (A) showed the distribution of GOLD BioAgeDiff in the UK Biobank (N = 417,067), with a mean of 0 and standard deviation of 5.11. Scatter density plot (B) showed a positive correlation of GOLD BioAgeDiff with chronological age (R = 0.225). Correlation of GOLD BioAgeDiff with health-related factors (C-E): disease counts (R = 0.12), Self-rated health (R = 0.24), and unhealthy lifestyles (R = 0.26).

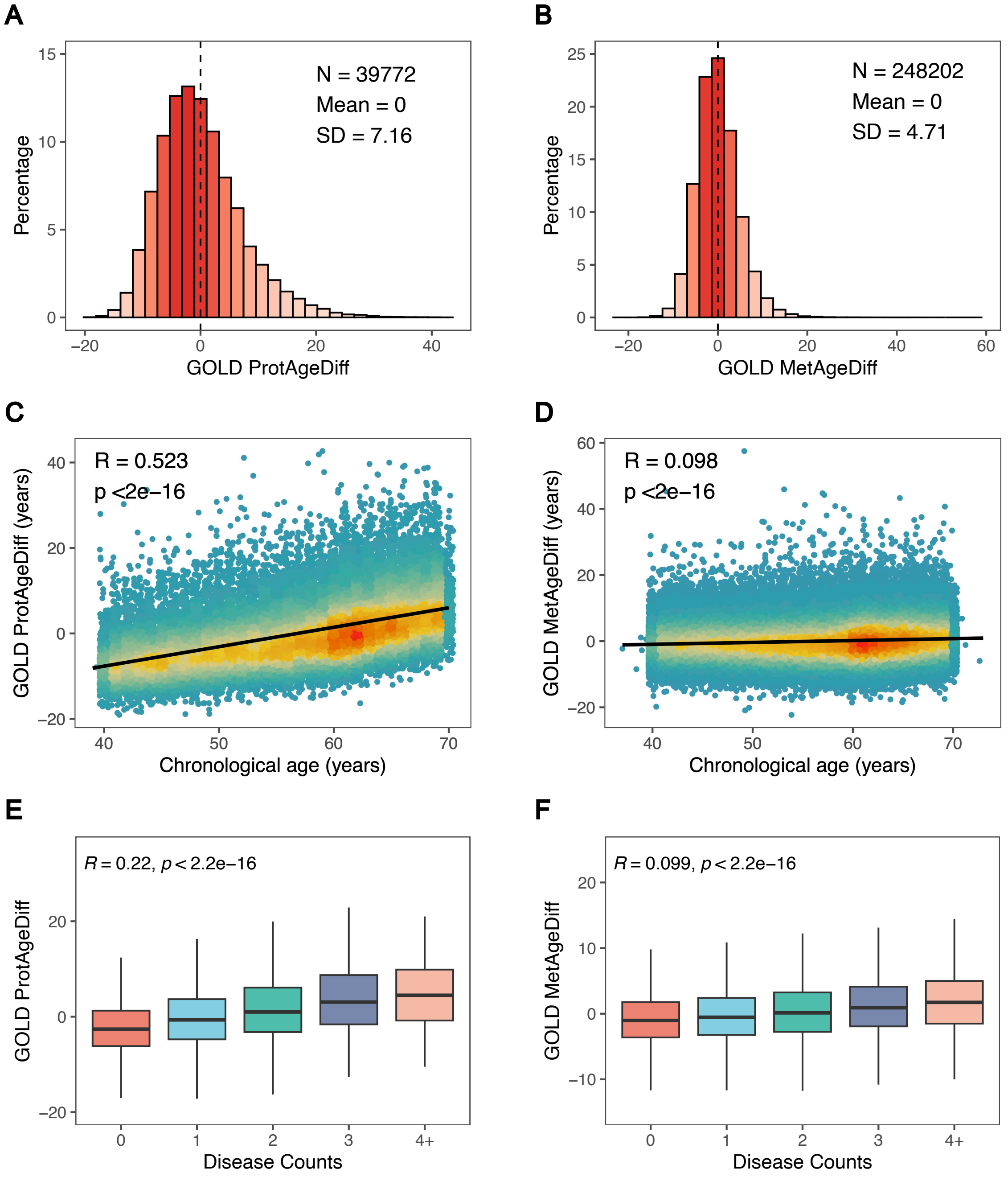


**Figure S4. GOLD ProtAgeDiff and MetAgeDiff in UKB.** The histogram (A) illustrated the distribution of GOLD BioAgeDiff in the UK Biobank dataset (N = 417,067), with a mean value of 0 and a standard deviation of 5.11. A scatter density plot (B) demonstrated a positive correlation between GOLD BioAgeDiff and chronological age (R = 0.225). The correlations of GOLD BioAgeDiff with various health-related factors were shown (C-E): disease counts (R = 0.12), self-rated health (R = 0.24), and unhealthy lifestyles (R = 0.26).

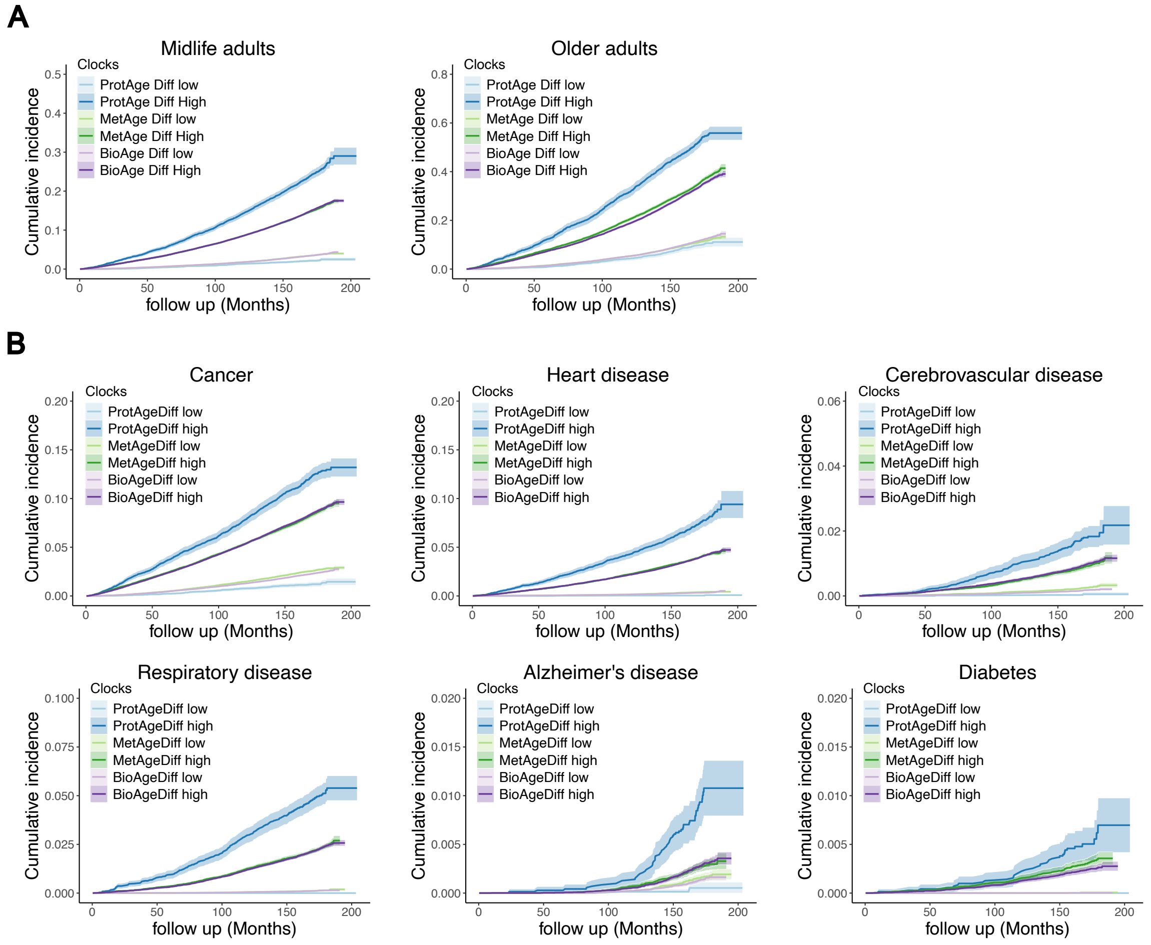


**Figure S5. Associations of ProtAgeDiff, MetAgeDiff, and BioAgeDiff with mortality in UKB.** The survival plots displayed the mortality trends of high-risk and low-risk groups over a follow-up period of approximately 16 years. The high-risk and low-risk categories were the top and bottom 20% of the differences of estimated biological age (ProtAge, MetAge and BioAge) and chronological age in the population. The cumulative incidences were investigated among midlife and older adults (A), and specific-cause mortality (caused by age-related chronic diseases) were also presented (B).

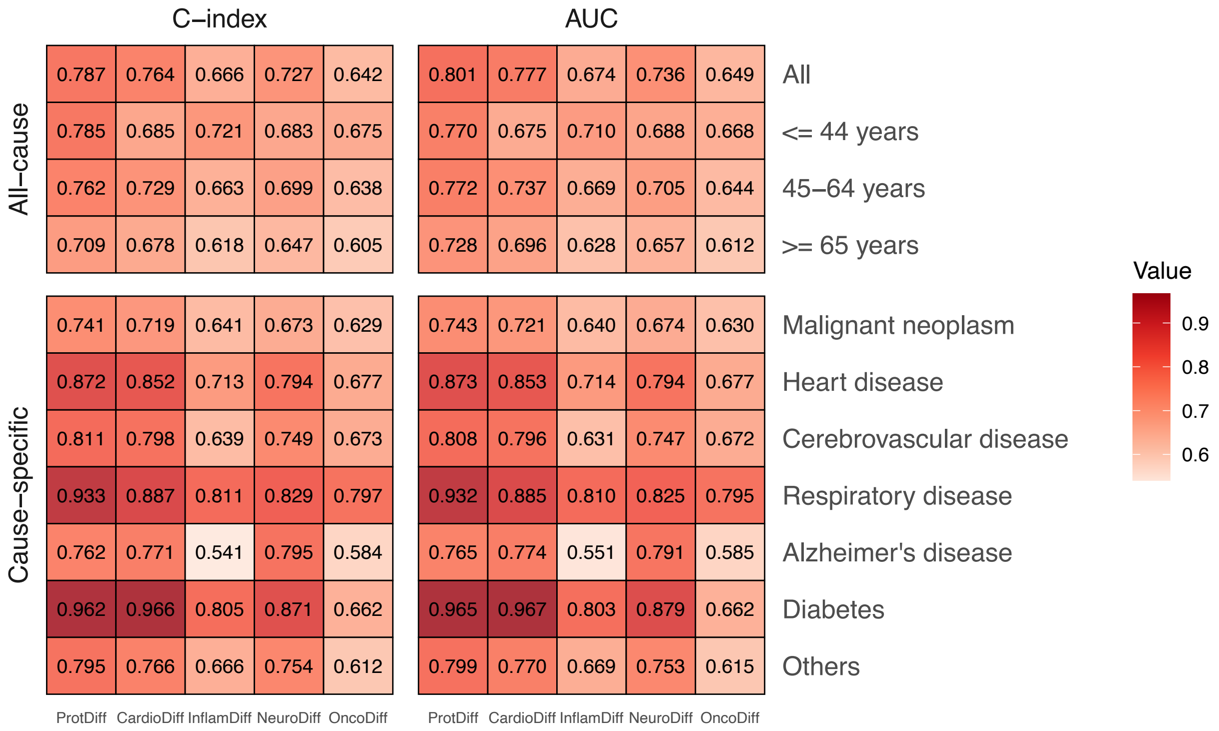


**Figure S6. The predictive capability of ProtAge and its subpanels regarding mortality.** The analysis included C-index in survival analysis and AUC value of 10-year mortality prediction for all-cause (age-stratified) and cause-specific mortality. The highest values were highlighted in bold. The terms ProtDiff, CardioDiff, InflamDiff, NeuroDiff, and OncDiff represented the composite scores of all, cardiometabolic, inflammation, neurology, and oncology proteins utilized in ProtAge.

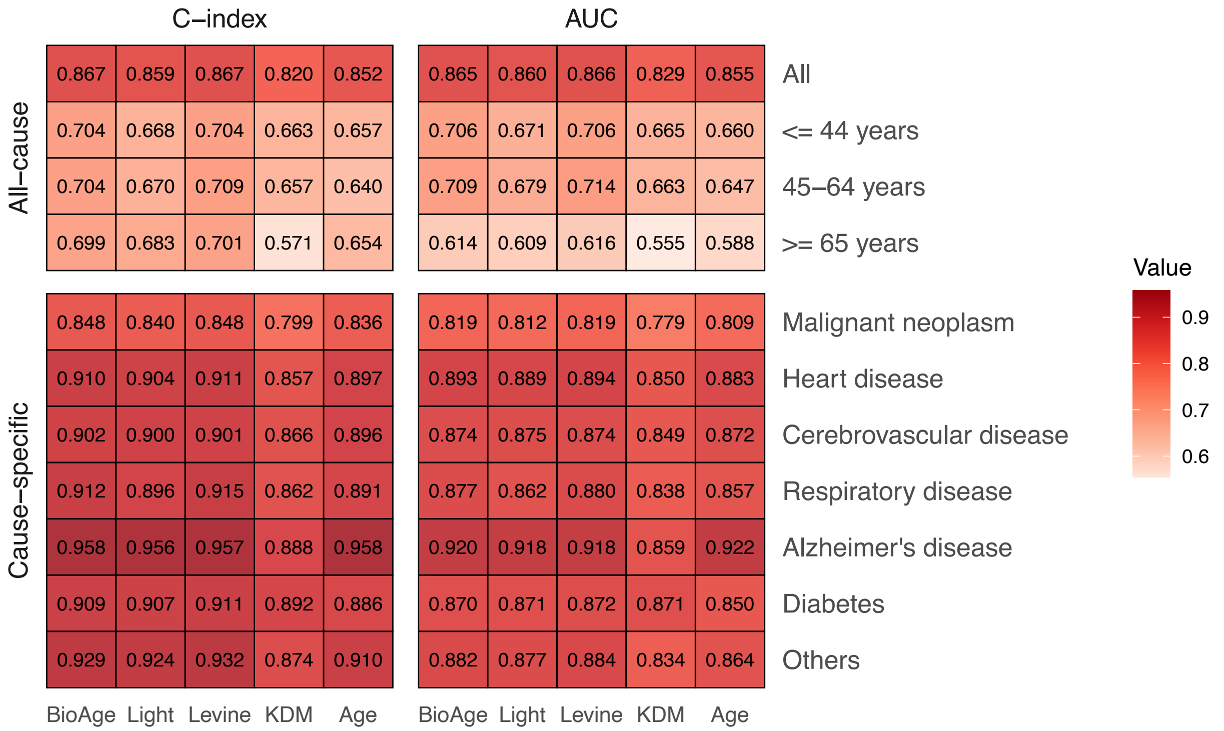


**Figure S7. The comparison of GOLD BioAge, and other common aging clocks in predicting mortality in NHANES III.** The evaluation included the C-index in survival analysis and the AUC value for predicting 10-year mortality using these aging clocks and chronological age for both all-cause (stratified by age) and cause-specific mortality. The highest values were denoted in bold. The BioAge, Light, Levine, and KDM referred to GOLD BioAge, its lighter version, Levine's phenotypic age, and the age derived from the KDM algorithm, respectively.

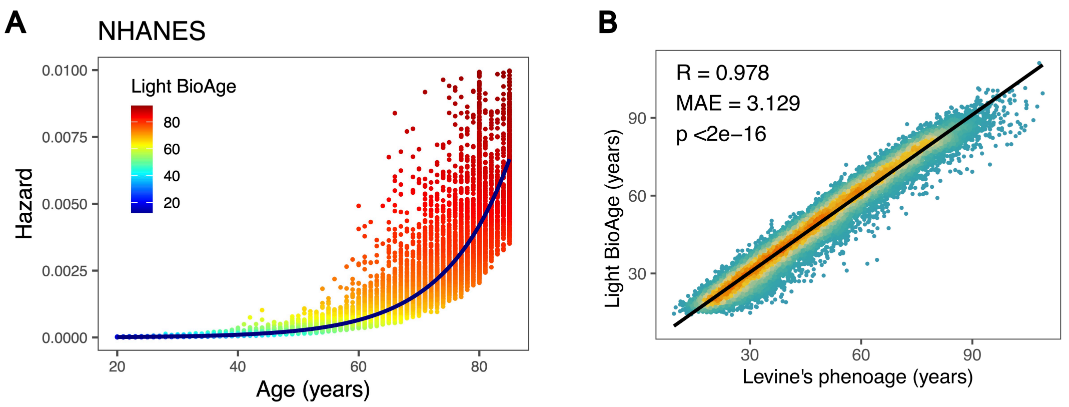


**Figure S8. The mortality hazard, Light BioAge and Levine's phenotypic age in NHANES.** Similar to GOLD BioAge, Light BioAge (A) indicated the age on the exponentially increasing line (Model 1) that corresponded with the mortality hazard of the colorful points (Model 2). The scatter density plot (B) revealed a strong correlation between Light BioAge and Levine's phenotypic age, demonstrating its validity as a simple measurement of biological aging.
