## Supplementary tables for "Gompertz law based biological age (GOLD BioAge): a simple and practical measurement of biological aging to capture morbidity and mortality risks"

**Table S1.** Information of 26 involved clinical biomarkers for feature selection to construct GOLD BioAge in NHANES.

**Table S2.** Information of 10 involved clinical biomarkers for feature selection to construct Light BioAge in NHANES.

**Table S3.** The statistic information of 10 indicators in formula of GOLD BioAge in NHANES and UKB.

**Table S4.** The statistic information of 4 indicators in formula of Light BioAge in five cohorts.

**Table S5.** Coefficients in the formula of GOLD BioAge.

**Table S6.** Coefficients in the formula of Light BioAge.

**Table S7.** Coefficients in the formula of GOLD ProtAge.

**Table S8.** Coefficients in the formula of GOLD MetAge.

**Table S9.** The associations of GOLD BioAge with mortality in NHANES and UKB.

**Table S10.** The associations of Light BioAge with mortality in NHANES and UKB.

**Table S11.** The associations of GOLD ProtAge and MetAge with mortality in UKB.

**Table S12.** The 31 items used to assess the frailty index in CHARLS.

**Table S13.** The sample sizes of GOLD aging clocks by disease counts, unhealthy lifestyles, and self-rated health.

**Table S1.** Information of 26 involved clinical biomarkers for feature selection to construct GOLD BioAge in NHANES.

| Biomarkers | Description | Biomarkers | Description |
|---|---|---|---|
| BMI | Blood mass index, kg/m^2 | RBC | Red blood cell count, 10^12 cells/L |
| SBP | Systolic blood pressure, mmHg | RDW | Red cell distribution width, % |
| DBP | Diastolic blood pressure, mmHg | MCV | Mean cell volume, fL |
| MBP | Mean blood pressure, mmHg | LogCRP | Log (c-reactive protein), mg/dL |
| GLU | Serum glucose, mmol/L | LYM% | Lymphocyte percent, % |
| HbA1C | Glycohemoglobin, % | NEU% | Neutrophils percent, % |
| HDL | High-density lipoprotein cholesterol, mg/dL | EOS% | Eosinophils percent, % |
| LDL | Low-density lipoprotein cholesterol, mg/dL | BASO% | Basophils percent, % |
| CHOL | Total cholesterol, mg/dL | MONO% | Monocyte percent, % |
| ALB | Albumin, g/dL | WBC | White blood cell count, 10^9 cells/L |
| ALP | Alkaline phosphatase, U/L | BUN | Blood urea nitrogen, mg/dL |
| TBIL | Total bilirubin, mg/dL | CREA | Creatinine, mg/dL |
| GGT | Gamma glutamyl transferase, U/L | UA | Uric acid, mg/dL |

**Table S2.** Information of 10 involved clinical biomarkers for feature selection to construct Light BioAge in NHANES.

| Biomarkers | Description | Biomarkers | Description |
|---|---|---|---|
| GLU | Serum glucose, mmol/L | LogCRP | Log (c-reactive protein), mg/dL |
| HbA1C | Glycohemoglobin, % | WBC | White blood cell count, 10^9 cells/L |
| HDL | High-density lipoprotein cholesterol, mg/dL | UA | Uric acid, mg/dL |
| CHOL | Total cholesterol, mg/dL | BUN | Blood urea nitrogen, mg/dL |
| RBC | Red blood cell count, 10^12 cells/L | CREA | Creatinine, mg/dL |

**Table S3.** The statistic information of 10 indicators in formula of GOLD BioAge in NHANES and UKB.

| Indicators | Units | NHANES (N = 39,348) | UKB (N = 417,067) |
|---|---|---|---|
| age | years | 49.50 (18.0) | 56.55 (8.09) |
| CREA | mg/dL | 0.88 (0.26) | 0.81 (0.16) |
| GLU | mmol/L | 5.56 (1.64) | 5.00 (0.62) |
| MCV | fL | 89.33 (5.70) | 91.18 (4.20) |
| RDW | % | 13.21 (1.20) | 13.44 (0.75) |
| ALB | g/dL | 4.22 (0.36) | 4.52 (0.26) |
| ALP | U/L | 69.81 (21.94) | 82.86 (21.88) |
| LYM% | % | 30.44 (8.54) | 28.84 (7.22) |
| WBC | 1000 cells/uL | 7.24 (2.11) | 6.84 (1.68) |
| GGT | U/L | 27.07 (24.98) | 32.63 (18.58) |

The mean (standard deviation) of biomarkers were shown in the table.

**Table S4.** The statistic information of 4 indicators in formula of Light BioAge in five cohorts.

| Indicators | Units | NHANES (N=38,001) | UKB (N=428,266) | |
|---|---|---|---|---|
| age | years | 49.72 (18.34) | 56.55 (8.09) | |
| CREA | mg/dL | 0.88 (0.26) | 0.81 (0.16) | |
| GLU | mmol/L | 5.48 (1.59) | 5.00 (0.62) | |
| LogCRP | mg/dL | 0.29 (0.292) | 0.98 (0.61) | |
| **Validation cohorts** | **Units** | **CHARLS (N=17,163)** | **CLHLS (N=2,546)** | **RuLAS (N=1,823)** |
| age | years | 55.90 (10.21) | 85.58 (12.06) | 77.02 (4.27) |
| CREA | mg/dL | 0.84 (0.29) | 0.94 (0.33) | 0.64 (0.17) |
| GLU | mmol/L | 5.75 (1.96) | 5.36 (1.80) | 6.02 (1.12) |
| LogCRP | mg/dL | 1.00 (0.64) | 1.00 (0.78) | 1.24 (0.51) |

The mean (standard deviation) of biomarkers were shown in the table.

**Table S5.** Coefficients in the formula of GOLD BioAge.

| Variable | Category | Units | Coefficient |
|---|---|---|---|
| age | | years | 1.0000 |
| CREA | Kidney | mg/dL | 5.2691 |
| GLU | Metabolic | mmol/L | 0.5797 |
| MCV | Erythrocyte | fL | 0.3389 |
| RDW | Erythrocyte | % | 2.6445 |
| ALB | Liver | g/dL | -4.7358 |
| ALP | Liver | U/L | 0.026 |
| LYM% | Immune | % | 0.2032 |
| WBC | Immune | 1000 cells/uL | 0.4459 |
| GGT | Kidney | U/L | -0.0608 |
| Constant ($\beta 0$) | | | -53.6287 |

**Table S6.** Coefficients in the formula of Light BioAge.

| Variable | Category | Units | Coefficient |
|---|---|---|---|
| age | | years | 1.0000 |
| CREA | Kidney | mg/dL | 8.3313 |
| GLU | Metabolic | mmol/L | 0.827 |
| LogCRP | Inflammation | mg/dL | 5.7305 |
| Constant ($\beta 0$) | | | -13.5298 |

**Table S7.** Coefficients in the formula of GOLD ProtAge.

| Variable | Category | UKB Code | Weight |
|---|---|---|---|
| Age | | | 1.0000 |
| ADGRG2 | Cardiometabolic | 57 | -0.6482 |
| EGFR | | 902 | -0.8457 |
| GDF15 | | 1137 | 1.3802 |
| IGFBP3 | | 1345 | -0.6716 |
| KIT | | 1507 | 0.0157 |
| LTBP2 | | 1631 | 1.6663 |
| NTproBNP | | 1912 | 0.7461 |
| REN | | 2270 | 0.7034 |
| AGER | Inflammation | 68 | -1.5969 |
| HGF | | 1264 | 1.3070 |
| LRRN1 | | 1622 | -0.5516 |
| PRSS8 | | 2161 | 0.9276 |
| BCAN | Neurology | 242 | -1.3198 |
| CDCP1 | | 483 | -0.1808 |
| NEFL | | 1840 | 1.1867 |
| TNFRSF10B | | 2717 | 0.4798 |
| ADGRG1 | Oncology | 56 | -0.6884 |
| CEACAM5 | | 512 | 1.4287 |
| EDA2R | | 883 | -0.8234 |
| HAVCR1 | | 1247 | 0.3592 |
| MMP12 | | 1733 | 0.4877 |
| WFDC2 | | 2891 | 1.2448 |
| Constant ($\beta 0$) | | | 0.0049 |

**Table S8.** Coefficients in the formula of GOLD MetAge.

| Variable | UKB Field ID | Weight |
|---|---|---|
| age | | 1.0000 |
| Free cholesterol to total lipids in small HDL percentage | 23647 | 0.5494 |
| Cholesteryl esters to total lipids in small LDL percentage | 23626 | -0.2878 |
| Cholesteryl esters to total lipids in large LDL percentage | 23616 | -0.1822 |
| Cholesteryl esters to total lipids in IDL percentage | 23611 | -0.3394 |
| Phospholipids to total lipids in very small VLDL percentage | 23604 | 0.8429 |
| Triglycerides to total lipids in large VLDL percentage | 23593 | -0.2614 |
| Cholesteryl esters in small HDL | 23576 | -0.543 |
| Triglycerides in chylomicrons and extremely large VLDL | 23487 | -1.1073 |
| Glycoprotein acetyls | 23480 | 2.4845 |
| Albumin | 23479 | -0.5125 |
| Creatinine | 23478 | 0.7854 |
| Acetone | 23477 | 0.4136 |
| Acetoacetate | 23476 | 0.4939 |
| 3-hydroxybutyrate | 23474 | -0.2927 |
| Lactate | 23471 | 0.2888 |
| Tyrosine | 23469 | 0.5285 |
| Phenylalanine | 23468 | 0.3799 |
| Valine | 23467 | -1.6374 |
| Glycine | 23462 | -0.8107 |
| Omega 6 fatty acids to omega 3 fatty acids ratio | 23459 | 0.6447 |
| Linoleic acid to total fatty acids percentage | 23456 | -1.8506 |
| Linoleic acid | 23449 | -0.3841 |
| Omega 3 fatty acids | 23444 | -1.7084 |
| Degree of unsaturation | 23443 | -1.0871 |
| Average diameter for VLDL particles | 23431 | -0.6324 |
| Glucose lactate | 20280 | 0.8090 |
| Constant ($\beta 0$) | | -0.6145 |

HDL: high-density lipoprotein cholesterol; LDL: low-density lipoprotein cholesterol; IDL: intermediate-density lipoprotein cholesterol; VLDL: very-low density lipoprotein cholesterol.

**Table S9.** The associations of GOLD BioAge with mortality in NHANES and UKB.

| **Category** | **Events** | **HR (95% CI)** | **P** |
|---|---|---|---|
| **NHANES (N = 39,348)** | | | |
| **All-cause mortality** | 4,716 | 1.091 (1.089-1.094) | <0.001 |
| <= 44 years | 275 | 1.085 (1.071-1.099) | <0.001 |
| 45-64 years | 993 | 1.114 (1.105-1.122) | <0.001 |
| >= 65 years | 3,448 | 1.102 (1.098-1.106) | <0.001 |
| **Cause-specific mortality** | | | |
| Cancer | 1,051 | 1.092 (1.087-1.097) | <0.001 |
| Heart disease | 1,231 | 1.122 (1.117-1.127) | <0.001 |
| Cerebrovascular disease | 264 | 1.125 (1.114-1.136) | <0.001 |
| Respiratory disease | 260 | 1.126 (1.115-1.137) | <0.001 |
| Alzheimer's disease | 167 | 1.155 (1.139-1.171) | <0.001 |
| Diabetes | 153 | 1.118 (1.104-1.132) | <0.001 |
| Other cause | 1,590 | 1.097 (1.093-1.101) | <0.001 |
| **UKB (N = 417,067)** | | | |
| **All-cause mortality** | 36,589 | 1.100 (1.099-1.101) | <0.001 |
| <= 44 years | 771 | 1.125 (1.111-1.139) | <0.001 |
| 45-64 years | 20,633 | 1.105 (1.103-1.107) | <0.001 |
| >= 65 years | 15,185 | 1.098 (1.095-1.101) | <0.001 |
| **Cause-specific mortality** | | | |
| Cancer | 21,299 | 1.087 (1.085-1.089) | <0.001 |
| Heart disease | 6,466 | 1.132 (1.128-1.136) | <0.001 |
| Cerebrovascular disease | 1,954 | 1.126 (1.120-1.133) | <0.001 |
| Respiratory disease | 3,200 | 1.165 (1.159-1.171) | <0.001 |
| Alzheimer's disease | 829 | 1.121 (1.111-1.131) | <0.001 |
| Diabetes | 313 | 1.158 (1.140-1.176) | <0.001 |
| Other cause | 10,439 | 1.112 (1.109-1.114) | <0.001 |

HR: hazard ratio; CI: confidence interval; All cox proportional hazard regression models were adjusted for age and sex.

**Table S10.** The associations of Light BioAge with mortality in NHANES and UKB.

| **Category** | **Events** | **HR (95% CI)** | **P** |
|---|---|---|---|
| **NHANES (N = 38,001)** | | | |
| **All-cause mortality** | 6,373 | 1.093 (1.091-1.095) | <0.001 |
| <= 44 years | 379 | 1.080 (1.066-1.095) | <0.001 |
| 45-64 years | 1,437 | 1.093 (1.085-1.102) | <0.001 |
| >= 65 years | 4,557 | 1.108 (1.103-1.112) | <0.001 |
| **Cause-specific mortality** | | | |
| Cancer | 1,402 | 1.097 (1.092-1.101) | <0.001 |
| Heart disease | 1,630 | 1.131 (1.126-1.136) | <0.001 |
| Cerebrovascular disease | 365 | 1.145 (1.134-1.157) | <0.001 |
| Respiratory disease | 352 | 1.133 (1.122-1.143) | <0.001 |
| Alzheimer's disease | 258 | 1.194 (1.178-1.210) | <0.001 |
| Diabetes | 205 | 1.127 (1.114-1.141) | <0.001 |
| Other cause | 2,161 | 1.102 (1.098-1.106) | <0.001 |
| **UKB (N = 428,266)** | | | |
| **All-cause mortality** | 37,619 | 1.102 (1.101-1.104) | <0.001 |
| <= 44 years | 802 | 1.113 (1.092-1.135) | <0.001 |
| 45-64 years | 21,192 | 1.101 (1.099-1.103) | <0.001 |
| >= 65 years | 15,625 | 1.102 (1.097-1.106) | <0.001 |
| **Cause-specific mortality** | | | |
| Cancer | 18,037 | 1.093 (1.091-1.095) | <0.001 |
| Heart disease | 5,475 | 1.130 (1.126-1.134) | <0.001 |
| Cerebrovascular disease | 1,667 | 1.134 (1.126-1.141) | <0.001 |
| Respiratory disease | 2,701 | 1.153 1.147-1.160) | <0.001 |
| Alzheimer's disease | 696 | 1.151 (1.139-1.164) | <0.001 |
| Diabetes | 262 | 1.158 (1.137-1.179) | <0.001 |
| Other cause | 8,781 | 1.105 (1.102-1.108) | <0.001 |

HR: hazard ratio; CI: confidence interval; All cox proportional hazard regression models were adjusted for age and sex.

**Table S11.** The associations of GOLD ProtAge and MetAge with mortality in UKB.

| Category | Events | HR (95% CI) | P |
|---|---|---|---|
| **ProtAge (N = 39,772)** | | | |
| **All-cause mortality** | 4,332 | 1.100 (1.097-1.103) | <0.001 |
| <= 44 years | 74 | 1.173 (1.145-1.202) | <0.001 |
| 45-64 years | 2,277 | 1.111 (1.107-1.116) | <0.001 |
| >= 65 years | 1,981 | 1.113 (1.107-1.119) | <0.001 |
| **Cause-specific mortality** | | | |
| Cancer | 1,717 | 1.086 (1.081-1.091) | <0.001 |
| Heart disease | 672 | 1.144 (1.136-1.152) | <0.001 |
| Cerebrovascular disease | 201 | 1.126 (1.111-1.141) | <0.001 |
| Respiratory disease | 385 | 1.185 (1.174-1.197) | <0.001 |
| Alzheimer's disease | 107 | 1.126 (1.105-1.147) | <0.001 |
| Diabetes | 39 | 1.218 (1.182-1.256) | <0.001 |
| Other cause | 1,211 | 1.107 (1.101-1.113) | <0.001 |
| **MetAge (N = 248,202)** | | | |
| **All-cause mortality** | 21,902 | 1.107 (1.105-1.108) | <0.001 |
| <= 44 years | 478 | 1.111 (1.093-1.129) | <0.001 |
| 45-64 years | 12,152 | 1.110 (1.108-1.113) | <0.001 |
| >= 65 years | 9,272 | 1.103 (1.100-1.107) | <0.001 |
| **Cause-specific mortality** | | | |
| Cancer | 10,506 | 1.094 (1.092-1.097) | <0.001 |
| Heart disease | 3,219 | 1.137 (1.133-1.142) | <0.001 |
| Cerebrovascular disease | 974 | 1.131 (1.123-1.139) | <0.001 |
| Respiratory disease | 1,624 | 1.163 (1.157-1.169) | <0.001 |
| Alzheimer's disease | 390 | 1.135 (1.122-1.148) | <0.001 |
| Diabetes | 169 | 1.194 (1.175-1.214) | <0.001 |
| Other cause | 5,020 | 1.118 (1.114-1.121) | <0.001 |

HR: hazard ratio; CI: confidence interval; All cox proportional hazard regression models were adjusted for age and sex.

**Table S12.** The 31 items used to assess the frailty index in CHARLS.

| No. | Description of the items | Metrics | Evaluation criteria |
|---|---|---|---|
| 1 | Self-reported physician diagnosed hypertension | | Yes = 1, No = 0 |
| 2 | Self-reported physician diagnosed diabetes | | Yes = 1, No = 0 |
| 3 | Self-reported physician diagnosed cancer | | Yes = 1, No = 0 |
| 4 | Self-reported physician diagnosed arthritis | | Yes = 1, No = 0 |
| 5 | Self-reported physician diagnosed chronic lung disease | | Yes = 1, No = 0 |
| 6 | Self-reported physician diagnosed liver disease | Age-related chronic diseases | Yes = 1, No = 0 |
| 7 | Self-reported physician diagnosed heart disease | | Yes = 1, No = 0 |
| 8 | Self-reported physician diagnosed cerebrovascular disease | | Yes = 1, No = 0 |
| 9 | Self-reported physician diagnosed any emotional, nervous, or psychiatric problems | | Yes = 1, No = 0 |
| 10 | Self-reported physician diagnosed memory-related disease | | Yes = 1, No = 0 |
| 11 | Self-reported vision problems | | Yes = 1, No = 0 |
| 12 | Self-reported hearing problems | | Yes = 1, No = 0 |
| 13 | Self-reported general health status | Self-rated health | Very poor or poor = 1, Very good, good, or fair = 0 |
| 14 | Difficulty with dressing | | Yes = 1, No = 0 |
| 15 | Difficulty with bathing or showering | | Yes = 1, No = 0 |
| 16 | Difficulty with eating | | Yes = 1, No = 0 |
| 17 | Difficulty with getting in and out of bed | | Yes = 1, No = 0 |
| 18 | Difficulty with using the toilet | Basic and instrumental activities of daily living | Yes = 1, No = 0 |
| 19 | Difficulty with controlling urination and defecation | | |
| 20 | Difficulty with doing household chores | | |
| 21 | Difficulty with managing money | | Yes = 1, No = 0 |
| 22 | Difficulty with taking medication | | Yes = 1, No = 0 |
| 23 | Difficulty with shopping for groceries | | Yes = 1, No = 0 |
| 24 | Difficulty with preparing meals | | Yes = 1, No = 0 |
| 25 | Mobility: difficulty with walking 100 yards or one block | | Yes = 1, No = 0 |
| 26 | Mobility: difficulty with getting up from a chair after sitting for long periods | | Yes = 1, No = 0 |
| 27 | Mobility: difficulty with climbing several flights of stairs without resting | Mobility capacity | Yes = 1, No = 0 |
| 28 | Mobility: difficulty with lifting or carrying weights over 10 pounds/jins | | Yes = 1, No = 0 |
| 29 | Mobility: difficulty with picking up a coin from the table | | Yes = 1, No = 0 |
| 30 | Mobility: difficulty with stooping, kneeling, or crouching | | Yes = 1, No = 0 |
| 31 | Mobility: difficulty with reaching arms above shoulder level | | Yes = 1, No = 0 |

Items 1-31 were categorized in to 0 or 1. The frailty index (FI) was calculated by the sum of the present health deficits divided by the total number of items (31 if no missing). The missing data was not included for the calculation. The frailty status was classified into three groups: robust (FI<=0.10), prefrail (0.1<FI<=0.25), and frail (FI>0.25).

**Table S13**. The sample sizes of GOLD aging clocks by disease counts, unhealthy lifestyles, and self-rated health.

| | GOLD BioAge | Light BioAge | GOLD MetAge | GOLD ProtAge |
|---|---|---|---|---|
| **NHANES** | | | | |
| **Disease counts** | **27, 587** | **27, 587** | | |
| 0 | 15, 340 | 15, 340 | | |
| 1 | 7, 253 | 7, 253 | | |
| 2 | 3, 503 | 3, 503 | | |
| 3 | 1, 173 | 1, 173 | | |
| 4+ | 318 | 318 | | |
| **Unhealthy lifestyles** | **5, 496** | **5, 527** | | |
| 0 | 78 | 78 | | |
| 1 | 583 | 584 | | |
| 2 | 1, 464 | 1, 472 | | |
| 3 | 1, 718 | 1, 727 | | |
| 4+ | 1, 653 | 1, 666 | | |
| **Self-rated Health** | 26, 034 | 26, 034 | | |
| Excellent or very good | 2, 528 | 2, 528 | | |
| Good | 7, 116 | 7, 116 | | |
| Fair | 10, 400 | 10, 400 | | |
| Poor | 5, 990 | 5, 990 | | |
| **UKB** | | | | |
| **Disease counts** | 270, 226 | 277, 431 | 160, 743 | 25, 457 |
| 0 | 172, 373 | 176, 975 | 102, 277 | 16, 108 |
| 1 | 74, 874 | 76, 871 | 44, 637 | 7, 093 |
| 2 | 18, 776 | 19, 276 | 11, 315 | 1, 834 |
| 3 | 3, 607 | 3, 698 | 2, 154 | 362 |
| 4+ | 596 | 611 | 360 | 60 |
| **Unhealthy lifestyles** | 307, 519 | 315, 697 | 182, 964 | 28, 942 |
| 0 | 3, 746 | 3, 867 | 2, 173 | 327 |
| 1 | 31, 720 | 32, 523 | 18, 220 | 2, 834 |
| 2 | 75, 054 | 77, 149 | 44, 202 | 7, 256 |
| 3 | 96, 519 | 99, 096 | 57, 762 | 9, 087 |
| 4+ | 100, 480 | 103, 062 | 60, 607 | 9, 438 |
| **Self-rated Health (total)** | 417, 067 | 425 ,679 | 246, 734 | 39, 484 |
| Excellent | 68, 432 | 70, 157 | 39, 657 | 6, 320 |
| Good | 240, 389 | 246, 795 | 143, 353 | 22, 468 |
| Fair | 87, 219 | 89, 667 | 52, 469 | 8, 623 |
| Poor | 18, 529 | 19, 060 | 11, 255 | 2, 073 |
