## Supplementary methods for "Gompertz law based biological age (GOLD BioAge): a simple and practical measurement of biological aging to capture morbidity and mortality risks"

**Derivation of the GOLD BioAge through Gompertz model**

The Gompertz regression, widely utilized for modeling mortality data, is primarily parameterized as a proportional hazards model. The Gompertz distribution is the two-parameter function with shape parameter $a$ and rate parameter $b$*^1^*. Its probability density function and hazard function are as follows:

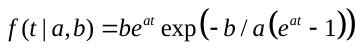

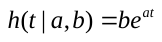

The hazard is increasing when shape $a>0$ and decreasing for $a<0$. For $a=0$ the Gompertz is equivalent to the exponential distribution with constant hazard and rate $b$. According to the Hazard function, the cumulative hazard model is

$$H\left( t \right)=b/a(e^{at}-1)$$

And the cumulative distribution function of the Gompertz model is

$$F\left( t|a,b \right)=1-exp(-b/a(e^{at}-1))$$

Then, we carried out two parametric proportional hazard model analysis with Gompertz distribution, one was regressed on chronological age (Model 1); another one was regressed on selected clinical biomarkers and chronological age to estimate the mortality risks (Model 2). The hazard functions were as followed:

*Model 1:* $h_{1}\left( t, age \right)=b_{1}exp\left( a_{1}t+{coef}_{1}[age] \right)$

*Model 2:* $h_{2}(t, age,biomarkers)=b_{2}exp(a_{2}t+{coef}_{2}[age,biomarkers])$

We defined the GOLD BioAge as the age reflecting the actual hazard risk. Therefore, we let the equation $h_{1}(t=0, GOLD BioAge)=h_{2}(t=0, age,biomarkers)$. Actually, the distributions of $h_{1}$ and $h_{2}$ were slightly different. The mean (standard deviation) of $h_{1}$ and $h_{2}$ were 8e-04 (0.0013) and 8e-04 (0.0013), respectively. We used a constant ($\gamma$) to correct the bias and let $h_{1}={\gamma*h}_{2}.$

And the GOLD BioAge was calculating as followed:

$$GOLD BioAge=\frac{1}{{coef}_{1}}(log(\frac{\gamma*b_{2}}{b_{1}})+{coef}_{2}\left[ age,biomarkers \right])$$

We further simplified the above formula. In the model 2, we fixed the coefficient of chronological age as that in model 1 (${coef}_{2}={coef}_{1}$). Then,

$$GOLD BioAge=age+ \frac{1}{{coef}_{1}}(log(\frac{\gamma*b_{2}}{b_{1}})+{coef}_{2}\left[ biomarkers \right])$$

**Estimate of the GOLD BioAge**

We applied a lasso penalized Cox regression model to select variables for constructing the Gompertz law based biological age (GOLD BioAge) using the NHANES training data. The mortality hazard was regressed on twenty-six clinical biomarkers and chronological age, with five-fold cross-validation to select the parameter value lambda (𝜆) for the penalized regression. Both the model performance and number of independent variables were considered to develop a sparse phenotypic age estimator. Finally, we chose lambda.1se, the value of 𝜆 that gave the most regularized model such that the cross-validated error was within one standard error of the minimum.

Consequently, the GOLD BioAge model involved 10 indicators, namely chronological age, creatinine, glucose, mean cell volume (MCV), red cell distribution width (RDW), albumin, alkaline phosphatase (ALP), lymphocyte percent (LYM), white blood cell count (WBC), gamma glutamyl transferase (GGT). The equation of GOLD BioAge was as followed:

$$GOLD BioAge=age +5.2691*creatinine +0.5797*glucose +0.3389*MCV$$

$$+2.6445*RDW-4.7358*albumin+0.0260*ALP$$

$$-0.2032*LYM +0.4459*WBC +0.0608*GGT-53.62873$$

In the UKB, we updated the coefficients, considering that the two population were different. The coefficients of the formula were as followed:

$$GOLD BioAge=age +5.3832*creatinine +1.4168*glucose+0.4206*MCV$$

$$+3.3162*RDW-5.0793*albumin+0.0385*ALP$$

$$-0.1899*LYM+0.9120*WBC+0.1007*GGT-78.6519$$

Moreover, for clinical practice simplicity, we developed a much lighter version (Light BioAge), which involved chronological age, creatine, glucose, and C-reactive protein (CRP). The Light BioAge was constructed using the following equation:

$$Light BioAge=age+8.3313*creatinine+0.8270*glucose$$

$$+5.7305*logCRP-13.5298$$

In the UKB, the formula of Light BioAge was as followed:

$$Light BioAge=age+9.0873*creatinine+1.7311*glucose$$

$$+4.1689*logCRP-20.1395$$

In the validations, we used the version of NHANES to calculate the Light BioAge in CHARLS, RuLAS and CLHLS cohorts.

**Comparison with Levine’s Phenotypic age**

Levine^2^ et al. estimates phenotypic age (Levine Phenotypic Age) reflecting the 10-year (t=120) cumulative mortality risk using the Gompertz cumulative distribution function. Notably, $F\left( t=120,PhenoAge \right)=10 year Mortality risk$. Two Gompertz proportional hazard models were constructed as followed:

$$F_{1}\left( t=120,PhenoAge \right)=1-exp(-exp(age*\overset{^}{\beta}) *(\exp(120*a)-1/a)$$

$$F_{2}\left( t=120,X \right)=1-exp(-exp(X\overset{^}{\beta}) *(\exp(120*a)-1/a)$$

where $\overset{^}{\lambda}=X\overset{^}{\beta}$ , by solved the equation $F_{1}(120, PhenoAge)=F_{2}(120, age,biomarkers)$. The Gompertz coefficients of $F_{1}$and $F_{2}$ were estimated separately.

The Phenotypic Age model is given by

$$Phenotypic Age=141.50225+\frac{ln\left( -0.00553*ln\left( 1-F_{2}(120,X) \right) \right)}{0.090165}$$

The mortality risk $F_{2}(120, X)$ was calculated through the Gompertz cumulative distribution function, using biomarkers and age. The linear combination form of Xβ was as followed:

$$X\beta=-19.907-0.0336*albumin+0.0095*creatinine +0.0195*glucose+0.0954*\log\left( CRP \right)-0.0120*LYM+0.0268*MCV+0.3356*RDW+0.00188*ALP+0.0554*WBC+0.0804*age$$

Considering that the Gompertz-based biological age (Levine Phenotypic Age^2,3^, Kuo’s proteomic age^4^, etc.) depended on the linear combination of involved variables (Xβ), we simplified the calculation process through the Gompertz hazard function. Both the 10-year mortality risk (cumulative distribution function) and mortality hazard (hazard function) were calculated by the Xβ. However, the mortality risk exhibited a dynamic change over 10 years, and the present mortality risk at baseline was more accurate. The calculation of GOLD BioAge showed the simplicity of our model, highlighting its efficiency to capture mortality risk through linear combination of clinical biomarkers.

**Assessment of health- related factors and outcomes**

(1) **Definition of Unhealthy Lifetyle Factors**

Unhealthy lifestyle score was based on six modifiable lifestyle factors: smoking, alcohol consumption, physical activity, diet, body mass index (BMI), and sedentary behavior. All these factors were defined consistent with World Health Organization^5-11^. They were obtained through structured questionnaires and 24-hour dietary recalls. Unhealthy BMI was defined lower than 18.5 kg/m^2^ or high than 24.9 kg/m^2^. Information on alcohol intake was self-reported. Low-risk alcohol assumption is determined as moderate drinking: no more than 2 drinks one day according to the dietary guidelines in the UK and US (one drink contains 8 g in the UK and 14 g ethanol in the US). In US NHANES, unhealthy smoke was defined as more than 100 cigarettes in a lifetime. We calculated the metabolic equivalent scores to define the physical activity. Participants were classified into thirds and the score lower than the top thirds of the population were defined as unhealthy people. Dietary quality was assessed by the Alternate Mediterranean Diet Score (aMED). We defined a healthy diet as the aMED higher than the median intake. The higher score indicated a healthier diet. Unhealthy sedentary behavior was defined as the duration higher than the upper quartile of distribution (Q4). In UK Biobank, unhealthy smoke was defined as no current smoking. Poor physical activity was defined as engaging in less than 75 minutes of vigorous activity or 150 minutes of moderate activity per week (or neither of these criteria were met), or failing to engage in vigorous activity at least once per week or moderate activity at least 5 days per week^6^. For diet, an high-risk level was defined as failing to meet the intake goals for 2 or more out of the following 3 components: a, total fruits and vegetables ≥ 4.5 pieces or servings per day (1 serving being 3 tablespoons; b, total fish intake ≥2 times per week; c, red meat intake: ≤ 5 times per week and processed meat intake ≤ 2 times per week. The detrimental sedentary behavior, following previous studies, were defined as the duration of computer and television (TV) watching time more than 3 hours per day. The participants were assigned scores ranging from 0 to 4, with 0 indicating no unhealthy lifestyles and 1, 2, 3, or 4 representing individuals with one or more unhealthy lifestyle factors. We explored the association between GOLD based biological age difference and unhealthy lifestyle score in NHANES 2007-2010 and UKB 2006-2010. For instance, in the UKB, the distribution of unhealthy lifestyle scores of GOLD BioAge was as follows: 0 (n = 3, 746), 1 (n = 31, 720), 2 (n = 75, 054), 3 (n = 96, 519), and 4 or more (n = 100, 480).

**Comorbidity**

We assessed the number of comorbidities, which included the following disease diagnoses. The count of comorbid conditions was categorized into five groups: no disease, 1 disease, 2 diseases, 3 diseases, and 4 or more diseases. In the NHANES dataset, the conditions included diabetes, high blood pressure, congestive heart failure, coronary heart disease, heart attack, stroke, cancer or malignancy, and chronic bronchitis. In the UK Biobank (UKB), the conditions considered were cancer, myocardial infarction, heart failure, stroke, chronic obstructive pulmonary disease (COPD), and dementia. For instance, in UKB, the distribution of GOLD BioAgeDiff, Light BioAgeDiff, MetAgeDiff, and ProtAgeDiff by comorbidity count was as follows: for GOLD BioAgeDiff (n = 270, 226): 0 (n = 172, 373), 1 (n = 74, 874), 2 (n = 18, 776), 3 (n = 3, 607), and 4+ (n = 596); for Light BioAgeDiff (n = 277, 431): 0 (n = 176, 975), 1 (n = 76, 871), 2 (n = 19, 276), 3 (n = 3, 698), and 4+ (n = 611); for GOLD MetAgeDiff (n = 160, 743): 0 (n = 102, 277), 1 (n = 44, 637), 2 (n = 11, 315), 3 (n = 2, 154), and 4+ (n = 360); for GOLD ProtAgeDiff (n = 25, 457): 0 (n = 16, 108), 1 (n = 7, 093), 2 (n = 1, 834), 3 (n = 362), and 4+ (n = 60).

(2) **Self-rated Health**

For self-rated health, there were four levels recorded across two cohorts: excellent or very good, good, fair and poor. For example, years of GOLD BioAgeDiff according to levels of self-rated health was subsequently categorized as excellent (n = 68,432), good (n = 240,389), fair (n = 87,219), poor (n = 18,529) among 417,067 participants in UKB.

**References**

1	Lee, E. T. & Wang, J. *Statistical methods for survival data analysis*. Vol. 476 (John Wiley & Sons, 2003).

2	Levine, M. E. *et al.* An epigenetic biomarker of aging for lifespan and healthspan. *Aging (Albany NY)* **10**, 573-591 (2018). <https://doi.org/10.18632/aging.101414>

3	Liu, Z. *et al.* A new aging measure captures morbidity and mortality risk across diverse subpopulations from NHANES IV: A cohort study. *PLoS Med* **15**, e1002718 (2018). <https://doi.org/10.1371/journal.pmed.1002718>

4	Kuo, C. L. *et al.* Proteomic aging clock (PAC) predicts age-related outcomes in middle-aged and older adults. *Aging Cell* **23**, e14195 (2024). <https://doi.org/10.1111/acel.14195>

5	Committee, D. G. A. *et al.* *Dietary guidelines for Americans 2015-2020*. (Government Printing Office, 2015).

6	Xie, H. *et al.* Association between healthy lifestyle and the occurrence of cardiometabolic multimorbidity in hypertensive patients: a prospective cohort study of UK Biobank. *Cardiovasc Diabetol* **21**, 199 (2022). <https://doi.org/10.1186/s12933-022-01632-3>

7	Zhan, Y., Yang, Z., Liu, Y., Zhan, F. & Lin, S. Interaction between rheumatoid arthritis and mediterranean diet on the risk of cardiovascular disease for the middle aged and elderly from National Health and Nutrition Examination Survey (NHANES). *BMC Public Health* **23**, 620 (2023). <https://doi.org/10.1186/s12889-023-15478-1>

8	Li, Y. *et al.* Trends in Self-Reported Adherence to Healthy Lifestyle Behaviors Among US Adults, 1999 to March 2020. *JAMA Netw Open* **6**, e2323584 (2023). <https://doi.org/10.1001/jamanetworkopen.2023.23584>

9	Li, X. *et al.* Accelerated aging mediates the associations of unhealthy lifestyles with cardiovascular disease, cancer, and mortality. *J Am Geriatr Soc* **72**, 181-193 (2024). <https://doi.org/10.1111/jgs.18611>

10	Zhang, Y. B. *et al.* Associations of healthy lifestyle and socioeconomic status with mortality and incident cardiovascular disease: two prospective cohort studies. *BMJ* **373**, n604 (2021). <https://doi.org/10.1136/bmj.n604>

11	Han, H. *et al.* Association of a Healthy Lifestyle With All-Cause and Cause-Specific Mortality Among Individuals With Type 2 Diabetes: A Prospective Study in UK Biobank. *Diabetes Care* **45**, 319-329 (2022). <https://doi.org/10.2337/dc21-1512>
